# Policies for Easing COVID-19 Pandemic Travel Restrictions

**DOI:** 10.1101/2021.04.14.21255465

**Authors:** Thien-Minh Le, Louis Raynal, Octavious Talbot, Hali Hambridge, Christopher Drovandi, Antonietta Mira, Kerrie Mengersen, Jukka-Pekka Onnela

## Abstract

During the COVID-19 pandemic, many countries implemented international travel restrictions that aimed to contain viral spread while still allowing necessary cross-border travel for social and economic reasons. The relative effectiveness of these approaches for controlling the pandemic has gone largely unstudied. Here we developed a flexible network meta-population model to compare the effectiveness of international travel policies, with a focus on evaluating the benefit of policy coordination. Because country-level epidemiological parameters are unknown, they need to be estimated from data; we accomplished this using approximate Bayesian computation, given the nature of our complex stochastic disease transmission model. Based on simulation and theoretical insights we find that, under our proposed policy, international airline travel may resume up to 58% of the pre-pandemic level with pandemic control comparable to that of a complete shutdown of all airline travel. Our results demonstrate that global coordination is necessary to allow for maximum travel with minimum effect on viral spread.

---

With more than 129 million cases and 2.8 million deaths globally as of March 31, 2021 (*1*), the COVID-19 pandemic has had an enormous impact on the world. The pandemic damaged the global economy, which shrank by 5.2% in 2020, the largest recession since World War II (*2*). With a patchwork of travel bans in place worldwide, the tourism industry has been severely affected, with estimated losses of 900 billion to 1.2 trillion USD and tourism down 58% to 78% (*3*). The airline industry has also suffered heavily, with 43 airlines declaring bankruptcy and 193 of 740 European airports at risk of closing (*4, 5*). To contain the pandemic, most countries took a two-pronged approach. First, they attempted to slow the spread of the disease internally by implementing various non-pharmacological interventions, such as social distancing, using face coverings, and closing businesses and schools. Second, they attempted to reduce the number of imported cases by implementing travel restrictions. While travel restrictions benefit the community by preventing importation of some infected cases, these policies end up costing the global economy an estimated 400 billion USD and millions of jobs each month (*6–8*). The gravity of the situation highlights the need for balance between protecting the health of the public and mitigating the short- and long-term economic damage related to infection control efforts.

The effectiveness of travel restrictions has been investigated in many studies (*9–15*) (see (*16–18*) for systematic reviews). Most of these studies suggest that travel restrictions are primarily effective at the early stage of a pandemic and may help to delay a pandemic up to 4–6 months (*11, 18*). However, the effect of travel restrictions wanes over time as cases are inevitably imported. Furthermore, the effect of travel restrictions is minimal relative to that of internal mitigation measures such as social distancing and mask wearing. Many researchers have concluded that continued use of travel restrictions is not worth the economic trade-off (*6, 17*). Although many studies have examined the effectiveness of travel restrictions, limited research has focused on the best way to lift these restrictions while still protecting health (*16*). Costantino et al. (2020) (*19*) and Linka et al. (2020) (*20*) studied partial removal of travel bans and urged caution for regions opening themselves up to regions with a more dire public health situation. Russell and colleagues went a step further by suggesting scenarios in which a country may want to leave travel restrictions in place (*21*). The authors argued that based on existing pandemic data and travel data, policymakers should first reconstruct the pandemic situation in each country and then estimate the number of imported cases they receive from each country. The ratio of imported cases to internal cases, together with the effective reproduction number, should then be used to decide whether travel restrictions are needed in that country. While these studies emphasize the important roles of imported and internal cases, none of them recommend specific strategies for easing travel restrictions or propose ways to coordinate them effectively to minimize health risks.

Our paper aims to address this gap in the literature. We developed a flexible network meta-population model for comparing the effectiveness of international travel policies, with a focus on evaluating the benefit of policy coordination. Because the epidemiological parameters of countries are unknown, they need to be estimated from data, a task usually accomplished using the likelihood function. However, complex stochastic models of infectious disease transmission often do not have computationally tractable likelihood functions. To overcome this limitation, we relied on a class of likelihood-free methods called approximate Bayesian computation (ABC). We then used our framework to examine two hypothetical travel-regulation policies that allow people to move from one country to another. The goal was to ensure that a country’s public health situation does not deteriorate after the country adopts the proposed travel policy. Theoretical results are provided to support the two proposed approaches. We also used simulation to compare the effectiveness of our recommended policies with existing travel restriction policies, such as a 14-day quarantine for all arrivals and a 14-day quarantine only for people returning from high-risk countries. Simulations indicate that our proposed travel policies would allow for more incoming travelers while maintaining control of the pandemic.

We considered a global travel model where people may travel from one country to another. In this network meta-population model, a node represents a country and an edge represents travel between two countries. The connections between the nodes are modeled using empirical travel data. To model the current state of the pandemic in each country, we used the epidemiological model presented by Warne et al. (*22*). In each country, at a given time, the population is divided into six mutually exclusive compartments: susceptible (*S*), undetected infected (*I*), active confirmed (*A*), confirmed recovered (*R*), confirmed deceased (*D*), and unconfirmed recovered (*R*^*u*^). Undetected infected (*I*) are individuals who have contracted COVID-19 but have not been identified; active confirmed (*A*) are individuals who have been identified as COVID-19 positive but are still receiving treatment or in self-quarantine; recovered confirmed (*R*) are individuals who have been confirmed to have recovered; and confirmed deceased (*D*) are individuals who were reported to have died from COVID-19. Recovered unconfirmed (*R*^*u*^) are individuals who have recovered from the disease but who were never confirmed as having contracted the virus. The remaining individuals in the population are susceptible (*S*) and could contract the virus. The spread of disease in each country evolves according to the following transitions and is governed by the indicated parameters:

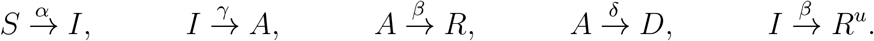

Here *α* is the transmission rate, *γ* is the identification rate, *β* is the recovery rate, and *δ* is the death rate.

Using this country-specific model, we then built a global network model of countries utilizing travel data as follows. In the meta-population network model, for a given country *i*, the status of its population is updated in two steps. First, the state of the epidemic evolves based on the internal population of country *i*. The transition from day *t* − 1 to *t* is characterized by the shift from **X**_*i*_(*t* − 1) based on the local, country-specific epidemiological model, where **X**_*i*_(*t* − 1) is a vector of six compartments of the status of country *i* at day *t* − 1. Second, the pandemic evolves based on factors external to each country; in this study, the external factor is travelers moving across borders.

In practice, the epidemiological model parameters for each country are unknown and need to be estimated from empirical data. Most statistical methods rely on the likelihood function for parameter estimation, but because our global model includes unobserved categories (susceptible (*S*), undetected infected (*I*), and unconfirmed recovered (*R*^*u*^)), we could not apply either frequentist or Bayesian inferential methods to this problem as both require a tractable likelihood function. Instead, we relied on a class of likelihood-free methods called approximate Bayesian computation (ABC). The use of ABC only requires the ability to forward simulate data from a model given model parameters; the corresponding likelihood function of the model does not need to be evaluated. In this paper, we used a variant method called replenishment ABC (RABC) (*23*). The main challenge of using ABC to calibrate our network meta-population model was the large number of parameters that needed to be estimated. Instead of using ABC to estimate all the parameters for all countries simultaneously, which is computationally expensive and may result in unstable parameter estimates, we used a marginal estimation strategy to estimate each country’s epidemiological parameters separately, while still taking the travel data into account. For a given country *i*, we first reconstructed all six states describing the pandemic situation in all other countries *j* ≠ *i* based on their epidemiological data. Based on the travel data, we then estimated the number of cases imported to country *i* from other countries. These quantities, together with the epidemiological data for country *i*, were then used to estimate the parameters for country *i*. More details on the estimation procedure are available in the Supplementary Materials.

Leaving borders completely open puts a country’s public health at risk, while closing borders is likely to have a negative effect on the economy. A policy that finds a middle ground between these two extremes is expected to provide a better balance between maintaining public health and preserving the economy. Some commonly used policies to ease travel restrictions include a 14-day quarantine for people traveling from high-risk regions and a 14-day quarantine requirement for all arrivals. However, there are no theoretical results demonstrating that these approaches control the pandemic as well as a full border closure would.

Under the 14-day quarantine for all arrivals policy, undetected infectious individuals transition to either active confirmed, recovered (confirmed or unconfirmed), or confirmed deceased as a result of monitoring during the quarantine period. As such, this policy helps stop importation of new undetected cases. However, this approach is also likely to dissuade travelers. Furthermore, if a large number of people are willing to travel despite the quarantine requirement, the country may see a surge in active confirmed cases from individuals undergoing mandatory quarantine. This surge could strain the receiving country’s healthcare system. To encourage travel, some countries have relaxed the quarantine requirement by dividing other countries into zones based on risk: travelers arriving from high-risk countries need to quarantine for 14 days whereas those arriving from low-risk countries have no quarantine requirement. While this approach could revitalize travel, it may still risk overburdening the receiving country’s healthcare system. Therefore, policy is needed that avoids these drawbacks and offers some guarantees that the pandemic remains under control.

In our global travel model, at the end of each day, we updated travel according to

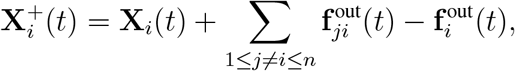

where 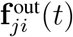 denotes the six compartments of individuals traveling from country *j* to country *i*. Our goal was to regulate the volume of inbound travel by only letting a certain proportion of travelers enter a country each day. We denote the proportions sequence 0 ≤ {*p*_*ji*_(*t*)}_1≤*j* ≠ *i*≤*n*_ ≤ 1 as the travel regulation sequence from country *j* to country *i* at day *t*, i.e., a temporal sequence of proportions of travelers permitted. A total shutdown of inbound travel in country *i* at day *t* is equivalent to *p*_*ji*_(*t*) = 0, ∀ 1 ≤ *j* ≠ *i* ≤ *n*, and fully open inbound travel in country *i* at day *t* is equivalent to *p*_*ji*_(*t*) = 1, ∀ 1 ≤ *j* ≠ *i* ≤ *n*. Under our strategy, at the end of day *t*, the status of country *i* is updated as follows:

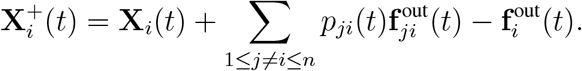

As a result, the number of undetected infected cases in country *i* at day *t* is also updated as 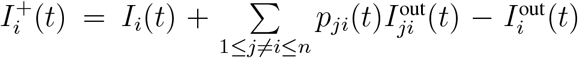. In our model, the undetected infected category is the only one that directly drives the epidemic. Therefore, if we can find a sequence {*p*_*ji*_(*t*)}_1≤*j* ≠ *i*≤*n*_ that ensures the number of undetected cases during the regulation period *T* does not go above a desired threshold *c*, then this sequence could be used to regulate travel. We took the following approach to find such a sequence. Consider a specific country with *I*(0) undetected cases initially, and suppose that under our regulation policy, we allow the number of daily undetected infected to be inflated at a rate *p*. In other words, if *I*(*t*) is the number of undetected cases evolved from the internal pandemic in a country at day *t*, then we allow incoming travel such that the number of undetected cases can increase up to *I*^+^(*t*) = *I*(*t*)(1 + *p*). Our goal is to find the value *p* so that the number of daily undetected infected cases during the regulation period stays below a given threshold *c*. Based on this value and the pandemic situation in the departure country, we can determine an appropriate sequence of proportions.

We considered two types of regulation. Regulation in terms of *average control* entails finding a proportion *p* such that the average number of daily undetected cases in the next *T* days stays below a fixed threshold *c*. Regulation in terms of *probability control* entails finding a proportion *p* such that the probability of daily undetected cases in the next *T* days staying lower than a threshold *c* is at least *π*. Lemma 1 and lemma 2 in the Supplementary Materials allowed us to find the proportion *p*.

Figure 2A shows the number of undetected infected cases in a country in the 7 days following the implementation of three different policies: fully open, fully closed, and our proposed average control policy with a threshold of *c* = 70. Note that the number of undetected cases under the average control scenario is below the required threshold and does not differ much from the one obtained under the fully closed scenario. Additionally, based on our calculations, the volume of inbound travelers under the average control policy can be up to 88.64% of the normal load. For more mathematical details on these calculations, including the proof, see the Supplementary Materials.

In practice, it may be hard to apply the proposed average control policies due to the logistical difficulties in regulation travel proportions daily. Therefore, we simplified these policies by first calculating the minimum value of the proportion sequence of incoming travelers. We then assigned the proportion of incoming travelers allowed as 0, 1*/*3, 1*/*2, or 1 if this minimum value belongs to ranges [0, 1*/*3), [1*/*3, 1*/*2), [1*/*2, 1), or [1, ∞), respectively.

We determined the effectiveness of six different travel regulation policies. We then compared their effectiveness in terms of the percentage of people allowed to travel relative to the pre-pandemic period. The first two policies were the most extreme: all countries are fully open or fully closed, denoted as policies P-1 and P-2, respectively. We investigated the effectiveness of the four remaining policies by having the receiving country adopt a given policy while all other countries remain fully open. Under P-3, the receiving country requires a 14-day quarantine for all arrivals. Under P-4, the country requires 14-day quarantine only for travelers from high-risk countries. A country is considered high risk if the average number of active confirmed daily cases exceeds 20 per 100,000 people in the last 2 weeks (*24*). Under P-5, the receiving country adopts the simplified version of the proposed average control policy, where travel is regulated such that the average number of daily undetected infected cases is at most 10% higher than the maximum number of daily cases under P-2. In P-6, the country adopts the simplified version of the proposed probability control policy, but travel is regulated such that the average number of daily undetected infected cases is at most 10% higher than the maximum daily cases under P-2 with a probability of at least 90%. Policy effectiveness was evaluated based on two factors: the percentage of inbound travelers and the epidemiological situation in the receiving country.

We evaluated inbound travel in two ways. The percentage of inbound travel capacity is the number of inbound travelers allowed under the policy divided by the number of inbound travelers under normal circumstances. The expected percentage of inbound travel is an adjusted version of the percentage of inbound travel capacity; if the 14-day quarantine policy is applied to people departing a country, we assume that only 5% of travelers from this country are willing to travel. South Korea requires a 14-day quarantine for all arrivals, and data provided by the Korea Tourism Organization supports this 5% assumption (*25*). After this adjustment, the percentage of expected inbound travelers is obtained by dividing the number of expected inbound travelers by the number of inbound travelers under normal circumstances, which gives us insight into the effect of the 14-day quarantine requirement. We report the effectiveness of policies on the epidemiological situation in the receiving country using three factors: the relative change in cases, the relative change in confirmed cases, and the percent of travelers who will eventually move to the active confirmed category after arrival. Relative change in cases is the difference between number of cases (detected and undetected) at the end and at the beginning of the regulated period divided by the number of cases at the beginning of the period; similarly, relative change in confirmed cases is the difference in the number of detected cases at the end and at the beginning of the regulated period divided by the number of detected cases at the beginning of the period.

The percent of travelers who will eventually move to the active confirmed category after arrival is calculated by using the number of undetected travelers who were eventually confirmed as having COVID-19 divided by the total number of incoming travelers. In these simulation settings, we considered only four representative countries, where each country has a reproduction number in the following ranges: 0.47–0.9, 0.9–1, 1–1.1, and 1.1–6.47. We first generated 200 sets of parameter values and the corresponding data sets using those parameters. For each data set, we used ABC to estimate the parameters of each country and the initial conditions at the time the policy was implemented. We then simulated 1000 realizations under different travel policies to obtain the above measurements, and report the 2.5th and 97.5th percentiles of each. To give a fair assessment on the effectiveness of travel restrictions on the pandemic, we report the outputs by stratifying the countries into three groups: Group 1 consists of countries with the effective reproduction number *R*(*t*) less than 0.9, Group 2 consists of countries with *R*(*t*) between 0.9 and 1.1, and Group 3 consists of countries with *R*(*t*) greater than 1.1. Finally, for each group, we calculated the average over these 200 iterations and used it to compare the effectiveness of different policies. Detailed simulation settings and comprehensive outputs for the effectiveness of different policies can be found in the Supplementary Materials.

Table 1 demonstrates how the different policies affect travel and each receiving country’s pandemic situation. Overall, our proposed average control policy, P-5, performed best at balancing the number of travelers and health outcomes. For all groups, the number of expected inbound travelers was highest for P-1, followed by P-4, then P-5. To satisfy the requirements dictated by P-6, countries had to eliminate inbound travel, rendering this policy equivalent to P-2. Under P-3 and P-4, approximately 0.09% to 0.11% of inbound travelers of Group 1 and Group 2 would become active confirmed. Therefore, if a receiving country has limited healthcare resources, it may experience challenges adopting P-3 or P-4. In terms of health outcomes, we observed that changing policies for countries in Group 3 does not offer much benefit, as the relative change in cases and confirmed cases were quite similar regardless of whether these countries totally close or fully open their borders. Travel restrictions were very effective for countries in Groups 1 and 2, with clear distinctions in the country’s pandemic situation upon adoption of the different travel regulation policies.

**Table 1:** Shown are 2.5^th^ and 97.5^th^ percentiles of travel effects and health outcomes for policies P-1 through P-6 using estimated epidemiological parameters to simulate epidemic and travel data. G1, G2, and G3 denote countries in Group 1, 2, and 3, respectively. RU is the relative change in the number of cases (including detected and undetected); RA is the relative change in the number of cases that were confirmed; IA is the percentage of incoming travelers who will eventually move to the active confirmed category after arrival; Tc is the percentage of inbound travel capacity; and Te is the percentage of expected of inbound travel.

|  |  | P-1 | P-2 | P-3 | P-4 | P-5 | P-6 |
| --- | --- | --- | --- | --- | --- | --- | --- |
| G1 | RU | (2.53, 3.20) | (0.06, 0.27) | (0.64, 0.92) | (0.88, 1.26) | (0.06, 0.27) | (0.06, 0.26) |
|  | RA | (1.58, 2.14) | (0.08, 0.27) | (0.86, 1.15) | (0.99, 1.36) | (0.08, 0.27) | (0.08, 0.27) |
|  | IA | (0.09, 0.11) | (0.00, 0.00) | (0.09, 0.11) | (0.09, 0.11) | (0.00, 0.00) | (0.00, 0.00) |
|  | Tc | 100% | 0% | 100% | 100% | 34% | 0% |
|  | Te | 100% | 0% | 5% | 89% | 34% | 0% |
| G2 | RU | (1.50, 2.05) | (0.45, 0.84) | (0.63, 1.02) | (0.86, 1.32) | (0.46, 0.84) | (0.45, 0.84) |
|  | RA | (0.99, 1.37) | (0.37, 0.64) | (0.60, 0.90) | (0.71, 1.04) | (0.37, 0.64) | (0.36, 0.64) |
|  | IA | (0.09, 0.11) | (0.00, 0.00) | (0.09, 0.11) | (0.09, 0.11) | (0.00, 0.00) | (0.00, 0.00) |
|  | Tc | 100% | 0% | 100% | 100% | 60% | 0% |
|  | Te | 100% | 0% | 5% | 89% | 60% | 0% |
| G3 | RU | (6.28, 6.65) | (6.30, 6.67) | (6.28, 6.65) | (6.28, 6.65) | (6.28, 6.65) | (6.28, 6.65) |
|  | RA | (5.32, 5.56) | (5.33, 5.57) | (5.32, 5.56) | (5.32, 5.56) | (5.32, 5.56) | (5.32, 5.56) |
|  | IA | (0.00, 0.00) | (0.00, 0.00) | (0.0, 0.00) | (0.00, 0.00) | (0.0, 0.00) | (0.00, 0.00) |
|  | Tc | 100% | 0% | 100% | 100% | 34% | 0% |
|  | Te | 100% | 0% | 5% | 100% | 34% | 0% |

To study the effectiveness of policy coordination, we investigated the percentage of people allowed to travel and the overall worldwide pandemic situation under different globally coordinated travel policies. Here we considered eight countries, where countries 1 and 2 have *R*(0) between 0.47 and 0.9, countries 3 and 4 have *R*(0) between 0.9 and 1, countries 5 and 6 have *R*(0) between 1 and 1.1, and countries 7 and 8 have *R*(0) between 1.1 and 6.47. The first two scenarios are the most extreme, where all countries are fully open or fully closed, denoted by S-1 and S-2, respectively. We used S-3 to denote the scenario where all countries require a 14-day quarantine for all arrivals; S-4 to denote the scenario where all countries use the simplified version of the average control policy; S-5 to denote the scenario where countries 1, 3, 5, and 7 require a 14-day quarantine for all arrivals while countries 2, 4, 6, and 8 are fully closed to inbound travel; and S-6 to denote the scenario where countries 1, 3, 5, and 7 use the simplified version of the proposed average control policy while countries 2, 4, 6, and 8 are fully closed to inbound travel. We used the same outcome measurements as in the previous simulation. Finally, we also evaluated the global coordination effectiveness by averaging the above measurements for all countries.

Table 2 reports the effectiveness of different coordinated responses in the three groups of countries along with a global average. Under S-4, where all countries use the proposed average control policy, the expected inbound travel increased up to 50% of normal travel, and the global pandemic situation was similar to that seen in scenarios where the borders are closed. These findings demonstrate that a global response is critical for containing the pandemic while maximizing safe travel.

**Table 2:** Shown are 2.5^th^ and 97.5^th^ percentiles of travel effects and health outcomes for scenarios S-1 through S-6 using estimated epidemiological parameters to simulate epidemic and travel data. See Table 1 caption for more information.

|  |  | S-1 | S-2 | S-3 | S-4 | S-5 | S-6 |
| --- | --- | --- | --- | --- | --- | --- | --- |
| G | RU | (10.68, 11.56) | (2.65, 3.06) | (4.02, 4.51) | (2.66, 3.07) | (3.45, 3.92) | (2.66, 3.07) |
|  | RA | (8.13, 8.89) | (2.77, 3.13) | (4.92, 5.43) | (2.77, 3.14) | (4.08, 4.55) | (2.77, 3.13) |
|  | IA | (1.57, 1.68) | (0.00, 0.00) | (1.57, 1.68) | (0.00, 0.01) | (0.80, 0.85) | (0.00, 0.00) |
|  | Tc | 100% | 0% | 100% | 50% | 50% | 25% |
|  | Te | 100% | 0% | 5% | 50% | 3% | 25% |
| G1 | RU | (11.16, 12.23) | (0.59, 0.93) | (3.17, 3.61) | (0.60, 0.94) | (1.84, 2.22) | (0.59, 0.93) |
|  | RA | (9.01, 9.95) | (0.74, 1.06) | (4.42, 4.90) | (0.75, 1.08) | (2.52, 2.91) | (0.75, 1.07) |
|  | IA | (1.98, 2.09) | (0.00, 0.00) | (1.98, 2.10) | (0.00, 0.01) | (1.00, 1.04) | (0.00, 0.00) |
|  | Tc | 100% | 0% | 100% | 64% | 50% | 32% |
|  | Te | 100% | 0% | 5% | 64% | 3% | 32% |
| G2 | RU | (12.13, 13.29) | (1.54, 2.13) | (2.98, 3.68) | (1.54, 2.13) | (2.50, 3.19) | (1.54, 2.13) |
|  | RA | (8.14, 9.14) | (1.62, 2.13) | (4.08, 4.81) | (1.62, 2.13) | (3.33, 4.04) | (1.62, 2.13) |
|  | IA | (1.77, 1.89) | (0.00, 0.00) | (1.77, 1.89) | (0.00, 0.01) | (0.89, 0.95) | (0.00, 0.00) |
|  | Tc | 100% | 0% | 100% | 64% | 50% | 32% |
|  | Te | 100% | 0% | 5% | 64% | 3% | 32% |
| G3 | RU | (7.31, 7.45) | (6.94, 7.08) | (6.94, 7.08) | (6.94, 7.08) | (6.95, 7.08) | (6.94, 7.08) |
|  | RA | (7.25, 7.35) | (7.10, 7.20) | (7.12, 7.22) | (7.10, 7.20) | (7.11, 7.21) | (7.10, 7.20) |
|  | IA | (0.77, 0.82) | (0.00, 0.00) | (0.77, 0.82) | (0.00, 0.00) | (0.42, 0.45) | (0.00, 0.00) |
|  | Tc | 100% | 0% | 100% | 7% | 50% | 3% |
|  | Te | 100% | 0% | 5% | 7% | 3% | 3% |

We used pandemic data from the Johns Hopkins University coronavirus data repository through May 31, 2020 (*26*). Flight data are from the Official Airline Guide (OAG). Because only data for January and February 2020 are available from OAG, we estimated flight data for other time periods using the OpenSky Network database (*27*). This database tracks the number of flights from one region to another over time, which can be used to calculate the rate of flight reduction and to estimate flight data for other months. We considered the starting day for each country to be the first day the country exceeded 500 total confirmed cases, because the estimation for the number of undetected infectious people is unstable during each country’s early pandemic period. For similar reasons, we only analyzed the 92 countries whose total number of confirmed infected cases exceeded 500 by April 15, 2020. The remaining countries were combined into a single fictional country labeled “Other.” All people traveling from this group of countries were assumed to be susceptible.

To demonstrate the ability of our model to capture the real evolution of the pandemic, we fit it to real data as follows. The fitting period starts when each country exceeded 500 total confirmed cases for the first time before May 31. The transmission rate *α* may change during the study. We first identify zero crossings of the second derivative of the function, i.e., where the function changes from convex to concave, and then use one value of *α* for the pre-period and another for the post-period. To reduce noise, we use a 7-day moving average, rather than daily values, to detect these change points. The initial condition for each country is chosen as (*S*(0), *I*(0), *A*(0), *R*(0), *D*(0), *R*^*u*^(0)), where *A*(0), *R*(0), *D*(0) are obtained from the real data, and assign *R*^*u*^(0) = 0. We obtain *I*(0) by simulating each country independently as in Warne et al. (2020) (*22*), where we allow *I*(0) to follow a uniform distribution with a range from 0 to 50 * *U* (0), where *U* (0) is the total confirmed cases in the country on that day. The median value of the posterior for *I*(0) is used as a point estimate for *I*(0). Due to reporting delays and data quality concerns, especially for the recovered confirmed cases, we only used the daily confirmed cases and daily deceased to construct the distance when performing the ABC algorithm to study the model’s parameters. The distance chosen for the calibration was the standardized Euclidean distance, defined as follows:

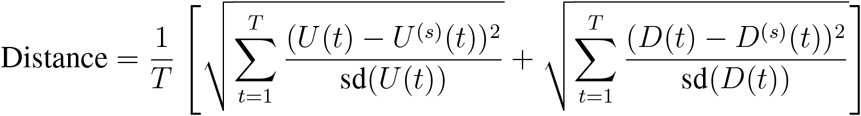

where *t* = 1, …, *T* are the days during the study period, *U (t*) is the total daily confirmed cases, *D*(*t*) is the daily deceased cases in that country at day *t*, and *U* ^(*s*)^(*t*) and *D*^(*s*)^(*t*) are the daily cases from simulated data. The prior standard deviations sd(*U (t*)) and sd(*D*(*t*)) were obtained by simulating the data for different combinations of parameters during the study period, and keeping 1000 realizations with the total number of confirmed and deceased cases at the end of the period no more than 5% different compared to the real data. To avoid the high rejection rate, we added one step by first running a preliminary analysis on each country independently and obtaining the posterior distribution for the parameters. We then used these parameters to simulate the data. To encourage more diversity in the realizations, we replaced the parameter *α* with *α* + runif(−*α/*2, *α/*2), where runif(−*α/*2, *α/*2) is a random number drawn from a uniform distribution from −*α/*2 to *α/*2. Based on these 1000 realizations, we then calculated sd(*D*(*t*)) and sd(*U (t*)) for *t* = 1, …, *T*.

Figure 1 shows the fit of our model for eight countries with the highest number of accumulated confirmed cases up to May 31, 2020: the United States of America (USA), Brazil (BRA), Russia (RUS), the United Kingdom (GBR), Spain (ESP), Italy (ITA), France (FRA), and India (IND). Overall, Figure 1 shows that our model effectively captures the real data (red), as the estimated line (blue) is very close to the observed line and is contained within the confidence interval.

**Figure 1:**
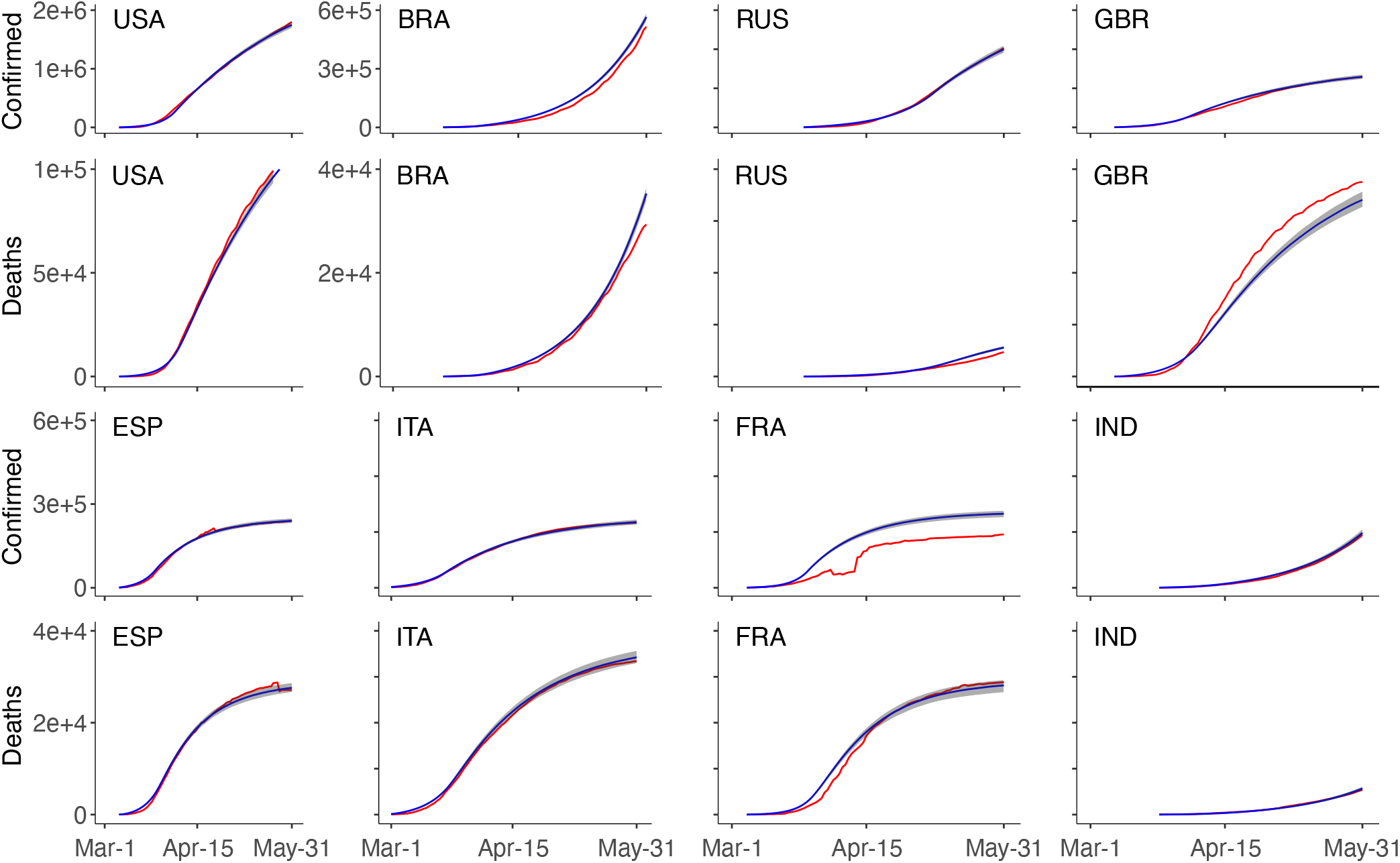
Model fit for different countries. For each country, the fit is demonstrated by the number of accumulated confirmed cases and the accumulated death confirmed. In each plot, the red line is the real data, the blue line is the median fitted values, and the shaded region is the 95% confidence interval. BRA, Brazil; ESP, Spain; FRA, France; GBR, Great Britain; IND, India; ITA, Italy, RUS, Russia; USA, United States.

To understand how different travel restriction policies affect the pandemic both globally and in each country, we used the estimated parameters for all countries before May 31 together with the travel data to simulate the course of the pandemic during the first 14 days of June 2020 under four different travel regulation scenarios. In the first scenario, we used 2020 travel data during the COVID-19 pandemic. In the second scenario, where all countries are fully open, we used 2019 travel data from the pre-pandemic period. In the third scenario, all countries fully closed their borders. Finally, we supposed all countries use the simplified average control policy.

Table 3 reports the relative change in the total number of cases, the total number of confirmed cases, and the inbound travel capacity for different countries. We evaluated the global effect of the pandemic for all countries and for the three groups of countries based on their *R*(*t*) values as defined earlier. The proposed simplified average control policy was the most effective in controlling the pandemic while still maximizing travel capacity. When countries used the proposed policy, the relative change in cases and confirmed cases was similar to those observed under the fully closed scenario. At the same time, the global travel rate remained as high as 58% compared to the fully open scenario. Additionally, the countries belonging to Group 1 benefited the most from travel restrictions with very little change in cases, even when comparing the most extreme scenarios. The 95% confidence interval for the relative change in cases for Group 1 was between 0.02 to 0.03 under the fully closed scenario and between 0.05 to 0.06 under the fully open scenario. Group 2 countries also saw only nominal benefit when closing the border compared to the fully open case. The 95% confidence interval for the relative change in cases decreased to (0.22, 0.26) under the fully closed scenario and to (0.24, 0.27) under the fully open scenario. Countries in Group 3 benefited the least from travel restrictions. The relative change in cases was between 0.80 to 0.84 under the fully closed scenario and between 0.81 to 0.85 under the fully open scenario. Finally, we also saw a huge reduction in global travel under the shutdown scenario with 2020 travel amounting to only 33% of 2019 travel.

**Table 3:**
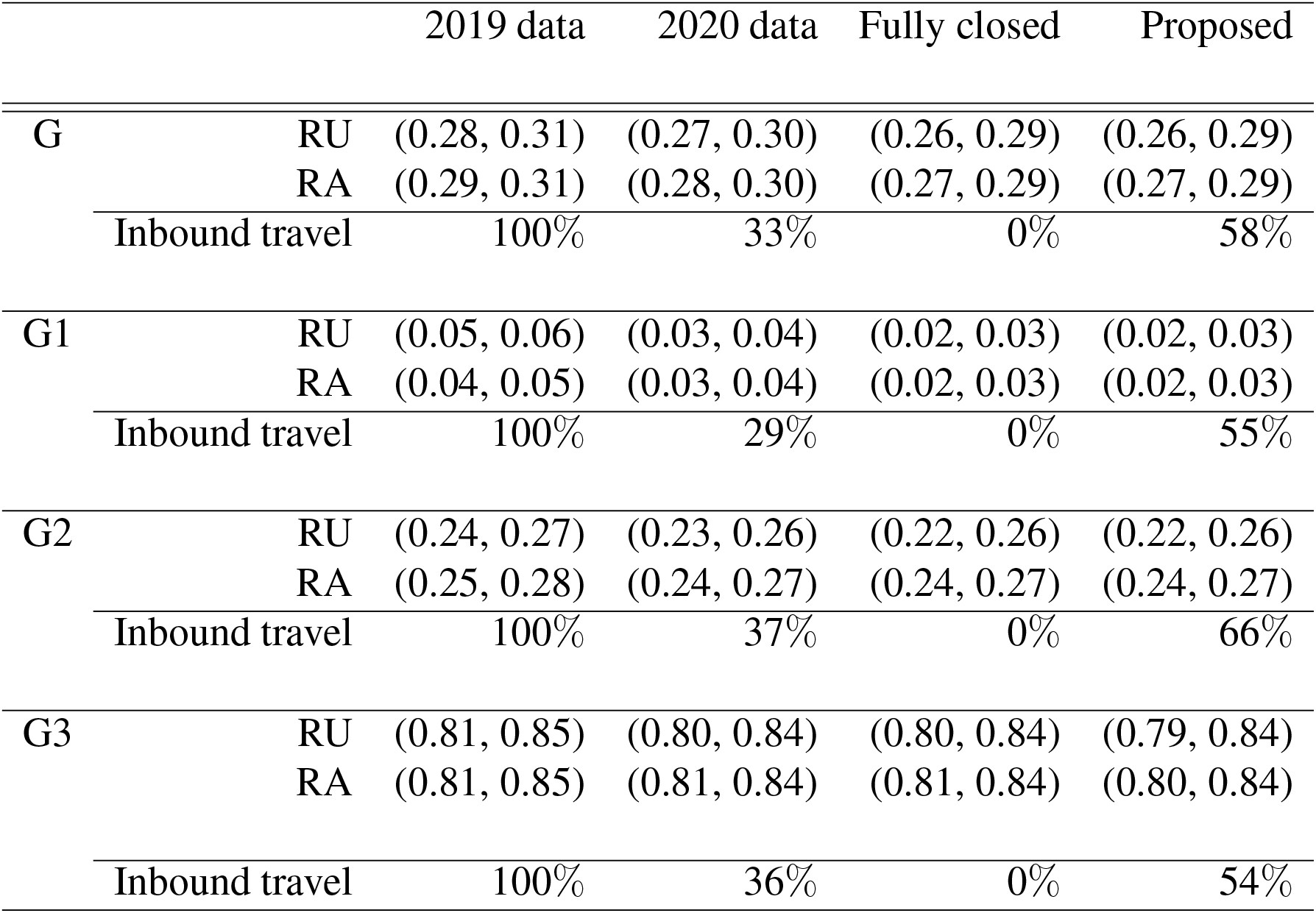
Shown are 2.5^th^ and 97.5^th^ percentiles of relative change in the pandemic situation and percentages of inbound travelers from different groups of countries for different opening scenarios. G denotes all countries; G1, G2, and G3 denotes countries in Group 1, 2, and 3, respectively. RelU is the relative change in the number of cases (including detected and undetected), and RelA is the relative change in the number of cases that were confirmed.

|  |  | 2019 data | 2020 data | Fully closed | Proposed |
| --- | --- | --- | --- | --- | --- |
| G | RU | (0.28, 0.31) | (0.27, 0.30) | (0.26, 0.29) | (0.26, 0.29) |
|  | RA | (0.29, 0.31) | (0.28, 0.30) | (0.27, 0.29) | (0.27, 0.29) |
|  | Inbound travel | 100% | 33% | 0% | 58% |
| G1 | RU | (0.05, 0.06) | (0.03, 0.04) | (0.02, 0.03) | (0.02, 0.03) |
|  | RA | (0.04, 0.05) | (0.03, 0.04) | (0.02, 0.03) | (0.02, 0.03) |
|  | Inbound travel | 100% | 29% | 0% | 55% |
| G2 | RU | (0.24, 0.27) | (0.23, 0.26) | (0.22, 0.26) | (0.22, 0.26) |
|  | RA | (0.25, 0.28) | (0.24, 0.27) | (0.24, 0.27) | (0.24, 0.27) |
|  | Inbound travel | 100% | 37% | 0% | 66% |
| G3 | RU | (0.81, 0.85) | (0.80, 0.84) | (0.80, 0.84) | (0.79, 0.84) |
|  | RA | (0.81, 0.85) | (0.81, 0.84) | (0.81, 0.84) | (0.80, 0.84) |
|  | Inbound travel | 100% | 36% | 0% | 54% |

Figure 2B demonstrates how much different countries benefited from travel restrictions during the first 2 weeks of June. Greece (GRC), Thailand (THA), Cyprus (CYP), and New Zealand (NZL) benefited most from border closure.

**Figure 2:**
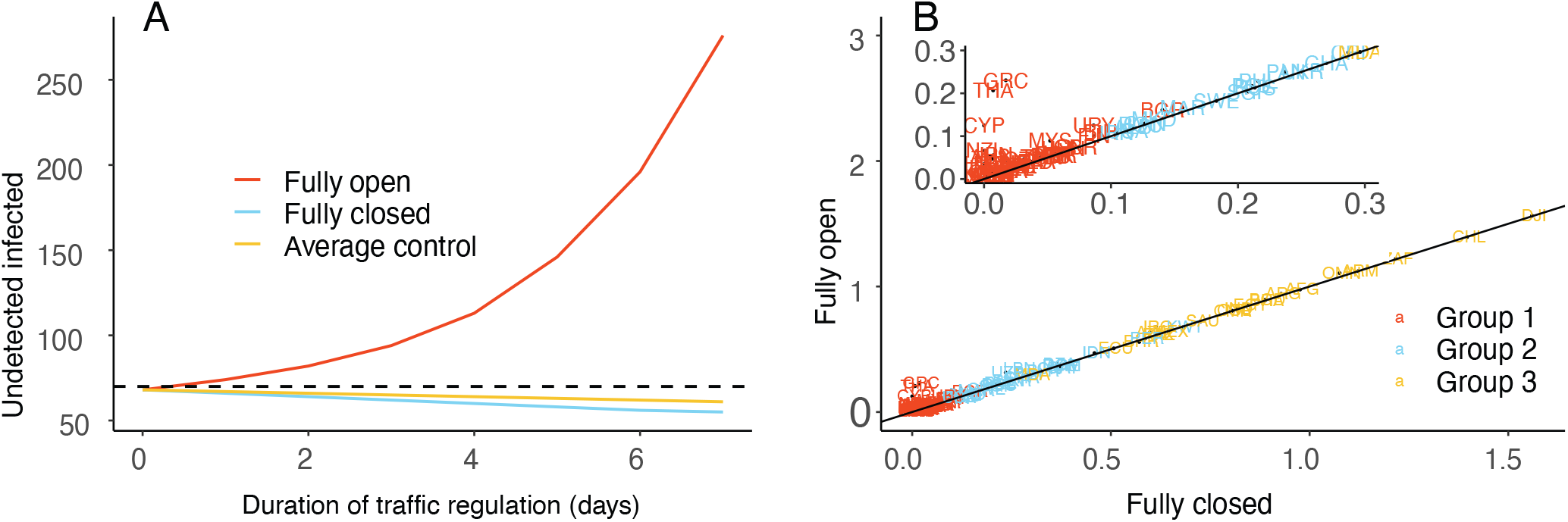
(**A**) Prediction of the average number of undetected infected cases for different travel regulation policies. “Fully open” indicates no travel restrictions are in place, “Fully closed” indicates no travel is permitted, and “Average control” denotes our proposed policy whereby the number of daily undetected infected cases stays below a threshold of *c* = 70 (the dashed line) on average. (**B**) Scatter plot for the relative change in total new cases for each country in the two most extreme scenarios: fully closed and fully open. The 97.5th percentile value of relative change in each country’s new cases under the fully closed scenario (x-axis) is plotted versus the fully open scenario (y-axis). The closer a country is to the reference line *x* = *y*, the less benefit that country gains from travel restrictions.

In this paper, we proposed a flexible network meta-population model for comparing the effectiveness of international travel policies and for assessing the benefit of international travel policy coordination. Using a mixture of simulation and theoretical findings, we showed that our proposed average control policies can effectively preserve global public health by reducing the number of cases while allowing international travel, thereby preserving the global economy. Our results show that globally coordinated travel policies are not only necessary for resuming international travel, but that it is also possible to accomplish this goal with minimal effect on public health relative to full border closure.

On the technical side, we proposed a marginal approach for estimating the epidemiological parameters for each country in a global network meta-population model. This approach helped overcome some of the difficulties of simultaneously or jointly estimating the model parameters.

Our statistical approach has one main limitation: we tried to control a hidden state of the model, the number of undetected infected cases *I*(*t*), which by definition is not available in the collected data. We therefore need to have a robust way to keep track of this number. Given the model, with the available data, we can approximate the hidden state *I*(*t*) by using the approximation 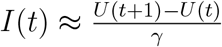, where *U (t*) = *A*(*t*) + *R*(*t*) + *D*(*t*) and *γ* is the identification rate that can be estimated from data. High-quality data is critical for tracking the number of undetected infected cases. In public health settings, one of the best strategies to estimate *I* is regular use of randomized serology testing (*28–30*).

Our model assumes active confirmed cases do not spread the disease. Since patients with COVID-19 may sometimes spread the disease to healthcare workers or their family members when they are in quarantine, this assumption may not hold in practice. Another limitation is that we used a conservative approach to model the global pandemic by assuming that travelers are either susceptible or undocumented infected. However, in reality, some travelers may be recovered confirmed or recovered unconfirmed cases, and therefore cannot infect anyone after arrival in another country. If this fact is taken into account, the number of people traveling may be higher than the currently reported numbers suggest. Unfortunately, the way empirical real data are currently reported does not reflect this fact.

Our proposed policies would allow travelers to enter a country without a quarantine requirement, with the travel rate adjusted based on the pandemic situation in the receiving country and the country of departure. Under this policy, the proportion of people allowed to travel from a high-risk country would be low, while a higher proportion of people would be permitted to travel from a low-risk country. Therefore, the proposed policy would allow control of the pandemic while encouraging travel, especially from low-risk countries.

The proposed travel regulation policies are designed for a meta-population model with local pandemic components as described in Warne et al. (2020) (*22*). Replacing this local infectious model (*22*) with different models such as SEIR or MSEIR as discussed in (*31*) can be easily accommodated by our modeling architecture. It is also possible to modify the proposed regulation policies to adapt to new virus variants.

Finally, although COVID-19 vaccines are becoming available and vaccination campaigns are underway at the time of writing, we have a long way to go before the entire world can achieve herd immunity, which is estimated to be attainable around 2024 (*32*), if ever (*33*). There is also increasing evidence that even as vaccines become more readily available, vaccine hesitancy remains high in some countries or some segments of the population (*34–37*). Also, with new variants of the virus emerging in South Africa, the U.K., and Brazil, countries are likely hesitant to lift their travel restrictions in the near future. For all of these reasons, evidence-based strategies that simultaneously preserve both global public health and the global economy are much needed.

## Data Availability

Proprietary flight data for January and February of 2020 are commercially available from the Official Airline Guide (OAG). The flight tracking data is publicly available from the OpenSky Network database https://opensky-network.org. The COVID-19 data is publicly available from the Johns Hopkins University coronavirus data repository https://github.com/CSSEGISandData/COVID-19. Reformatted data and R code used in this study are publicly available at https://github.com/onnela-lab/covid-travel.

## Funding

T.M.L., L.R., and J.P.O. were supported by NIH award AI138901 (Onnela). A.M. was partially supported by SNF grant 105218 163196.

## Author contributions

J.P.0., T.M.L., A.M., K.M., and C.D. designed the research; T.M.L. performed the research; T.M.L., L.R., and O.T. prepared the code and analyzed the data; T.M.L., J.P.0., H.H., L.R., C.D., O.T., A.M., and K.M. wrote and edited the paper.

## Competing interests

The authors declare no other relationships or activities that could appear to have influenced the submitted work.

## Supplementary Materials

Supplementary Materials can be found at the Supplementary Materials for Policies for Easing Pandemic Travel Restrictions. The Supplementary Materials include:

Models and Methods Simulation Studies

Figure S1

Tables S1-S7

References

## Supplementary Materials

### 1 Models and methods

#### 1.1 Local travel model

We first consider a local epidemiological model as in Warne et al. (2020) (*1*). In this local model, for each country, at a given time, its population’s status is divided into 6 mutually exclusive compartments: susceptible (*S*), undetected infected (*I*), active confirmed (*A*), confirmed recovered (*R*), confirmed deceased (*D*) and unconfirmed recovered (*R*^*u*^). Its dynamic states evolve as:

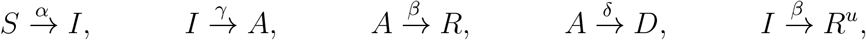

where *α* is the transmission rate, *γ* is the identification rate, *β* is the recovery rate, and *δ* is the death rate. Suppose that for a given country the status of its population at time *t* is **X**(*t*) = [*S*(*t*), *I*(*t*), *A*(*t*), *R*(*t*), *D*(*t*), *R*^*u*^(*t*)], and *θ* = (*α, β, δ, γ*) represents the parameter of the statistical model for the country. Using the tau leaping method by Gillespie (2001) (*2*), the status of its population at time (*t* + *τ*) evolves as 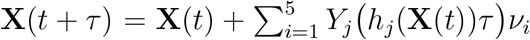. In the above formula, *ν*_*i*_, *i* = 1, …, 5, are the transition vectors, *ν*_1_ = [−1, 1, 0, 0, 0, 0]^*T*^, *ν*_2_ = [0, −1, 1, 0, 0, 0]^*T*^, *ν*_3_ = [0, 0, −1, 1, 0, 0]^*T*^, *ν*_4_ = [0, 0,−1, 0, 1, 0]^*T*^, and *ν*_5_ = [0, −1, 0, 0, 0, 1]^*T*^. Let the random variables *Y*_*i*_ (*h*_*i*_(**X**(*t*))*τ*) be Poisson distributed with rates *h*_*i*_(**X**(*t*)*τ*), for *i* ∈ {1, …, 5}. More specifically, 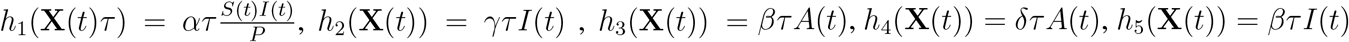, and *P* is the country’s population.

We choose *τ* = 1, which represents the change in population status after each day. Then the dynamic evolution of the epidemic in the country can be elaborated further as follows. After each day, the state of the model evolves from **X**(*t*) = [*S*(*t*), *I*(*t*), *A*(*t*), *R*(*t*), *D*(*t*), *R*^*u*^(*t*)] to **X**(*t* + 1) = [*S*(*t* + 1), *I*(*t* + 1), *A*(*t* + 1), *R*(*t* + 1), *D*(*t* + 1), *R*^*u*^(*t* + 1)] by the transformation 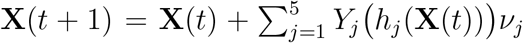. In particular, 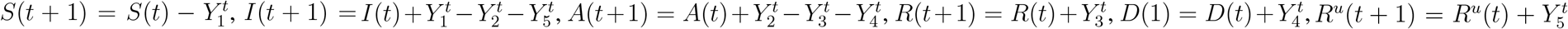, where 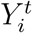 are Poisson distributed with rates *h*_*i*_(**X**(*t*)), *i* = 1, …, 5, 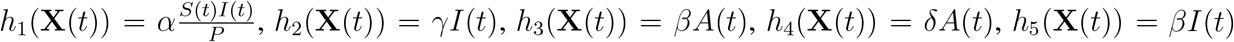.

Notice that the local model can be made more flexible by letting the transmission rate *α* change over time, i.e., setting *α* = *α*_1_ℐ_0,*T* (1)_(*t*) + *α*_2_ℐ_*T* (1),*T* (2)_(*t*) + …+ *α*_*m*_ℐ_*T (m−*1),*T (m*)_(*t*), where 0 = *T* (0) *< T* (1) *<* …*< T (m*) = *T*, and the indicator function ℐ_*T (i),T (i*+1)_(*t*) = 1 if *T (i) < t* ≤ *T (i* + 1), and 0 otherwise.

#### 1.2 Global travel model

Our global epidemiological model model is built based on the local model by utilizing travel flow data as follows. For a given country *i*, suppose the status of its population at the end of day (*t* − 1) is 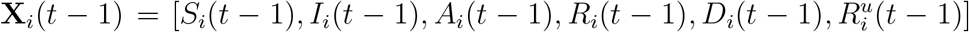, and the parameter of the statistical model for this country is *θ*_*i*_ = (*α*_*i*_, *β*_*i*_, *δ*_*i*_, *γ*_*i*_). On day *t*, the epidemic state in country *i* is updated via two steps. First, the state evolves based on country *i*’s internal population. Second, the state evolves based on external factors, here the inflow of airline travelers from other countries and the outflow of airline travelers to other countries.

We consider changes due to internal effects first. For country *i*, the transition from *t* − 1 to *t* is characterized by the shift from 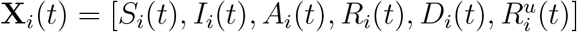 where

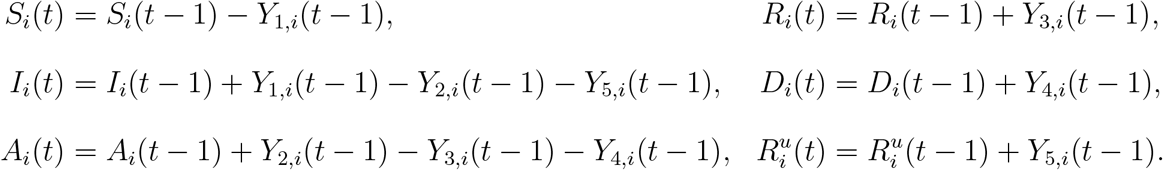

and *Y*_*j,i*_(*t* − 1), *j* = 1, …, 5, are Poisson distributed with rates

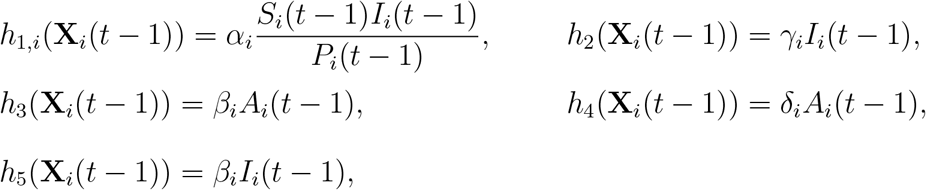

and *P*_*i*_(*t* − 1) is the size of the population in country *i* on day (*t* − 1).

The travel data specify how many new individuals enter the country on day *t* from each of the disease states. The current state is updated as 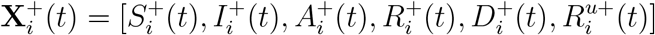, where 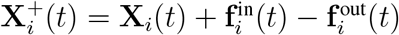, where 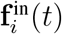 represents the six compartments of people entering the country on day *t* and 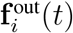 represents the six compartments of people leaving the country on day *t*. Due to temperature checks and other approaches for screening travelers, we assume that all active confirmed cases are unable to travel. We also assume that deceased individuals do not travel between countries. Consequently, the compartments in 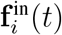 and 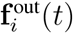 only include four of the six disease states: susceptible (*S*), undetected infected (*I*), recovered confirmed (*R*), and recovered unconfirmed (*R*^*u*^). Travelers in the recovered confirmed (*R*) and recovered unconfirmed (*R*^*u*^) states do not impact the epidemiological state of destination population. However, data on all four categories is not readily available. While each country keeps track of the total number of confirmed recovered each day, they do not necessarily keep track of how many of them leave the country. Therefore, we take a conservative approach and assume that each traveler either belongs to the *S* category or the *I* category, meaning travelers bring some potential risk when they arrive in a new country as undetected infected will likely spread the disease and susceptible individuals reduce population immunity and can proliferate disease spread. In other words, we impose 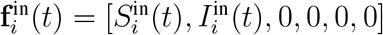 and 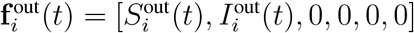, where 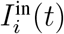 and 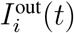 are the number of undetected infected that enter and leave country *i* on day *t*, respectively. 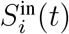 and 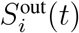 are the total numbers of susceptible individuals who enter and leave the country *i* on day *t*, respectively. 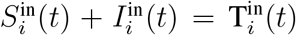 gives the total number of travelers that enter country *i* on day *t*, and 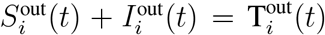 gives the total number of individuals who leave the country *i* on day *t*. As such, **X**_*i*_(*t*) and 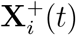 only differ in the first two categories, where 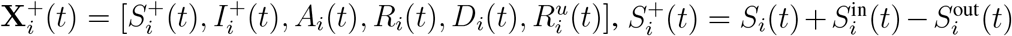, and 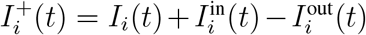. On day (*t* + 1), the internal model will be updated based on 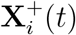. The compartmental quantities are updated as follows

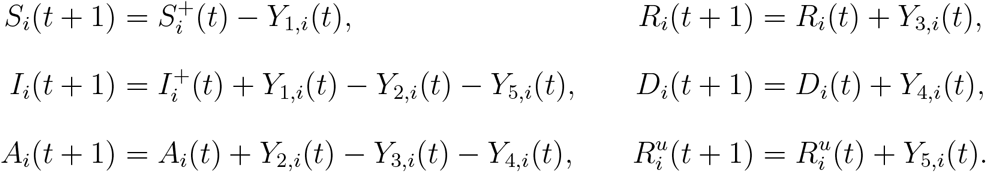

and *Y*_*j,i*_(*t*), *j* = 1, …, 5, are Poisson distributed with rates

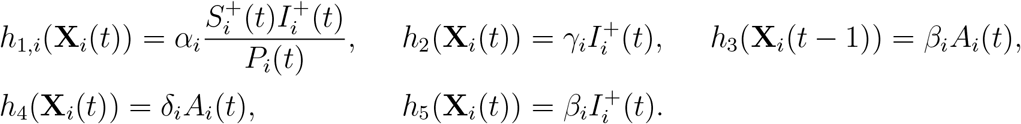

Our model assumes that active confirmed cases do not spread the disease due to self-isolation or hospitalization. Therefore, undetected infected cases are the only ones to spread the disease. Moreover, when *I* = 0, the pandemic in the country will cease if we stop admitting undetected infected cases from other countries. Each day, among the people that travel from country *i* to other countries, there may be some undetected infected cases. If an undetected infected individual enters a country with zero undetected infectious cases, they will seed a new outbreak in this country. Suppose that on day *t*, there are 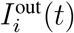 undetected infected people departing from country *i* to country *j, j ≠ i*. Then 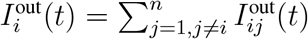, where 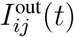 is the number of undetected infected moving from country *i* to country *j* at day *t*, and *n* is the total number of countries. We model the number of undetected infected people who are leaving country *i* for country *j* at day *t* using a multinomial distribution with probabilities based on travel network data. In other words, 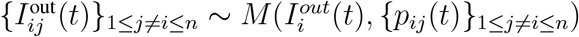, where 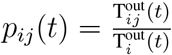 and 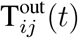 is the total number of travelers leaving country *i* for *j* at day *t*. Let us denote 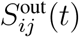 as the number of susceptible people who travel from country *i* to country *j* at day *t*. Then 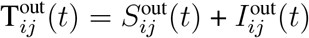. Therefore, at the end of day *t*, the six states for country *i* are updated as 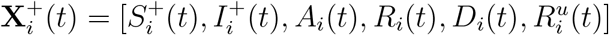, where

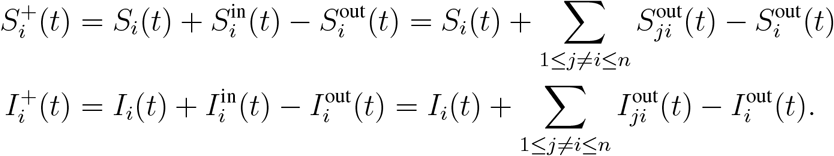

#### 1.3 Travel regulation policies

Our goal is to find the value *p* so that the number of undetected infected *I*^+^(1), *I*^+^(2), …, *I*^+^(*T*) stay below a given threshold *c*. More specifically, we consider two types of regulation, an average control policy and a probability control policy described below:

1. Regulation in terms of **average control**, where we find a proportion *p* such that the average number of undetected cases each day in the next *T* days stays below a threshold *c*, i.e., *E*(*I*^+^(1)), *E*(*I*^+^(2)), …, *E*(*I*^+^(*T*)) < *c*, where *E* denotes the expectation.
2. Regulation in terms of **probability control**, where we find a proportion *p* such that the probability of undetected cases each day in the next *T* days being lower than a threshold *c* is at least at *π*, i.e. *P (I*^+^(1) < *c, I*^+^(2) < *c*, …, *I*^+^(*T*)^+^ < *c*) ≥ *π*.

The following lemmas gives us the proportion *p* that satisfies the above requirements.

##### Lemma 1.

*Under the assumptions of our model, for a given country with population size P, the initial status* **X**(0) = [*S*(0), *I*(0), *A*(0), *R*(0), *D*(0), *R*^*u*^(0)] *and the parameter of the statistical model is θ* = (*α, β, δ, γ*), *let us denote* 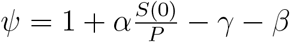,

1. *The average control requirement is satisfied if*

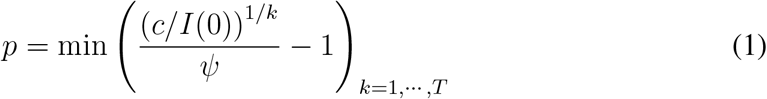
2. *The probability control requirement is satisfied if*

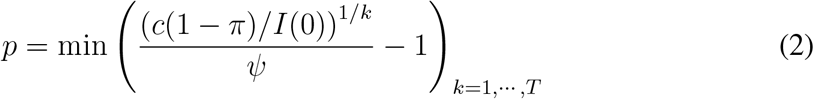

*Proof:*

1. *Average control. With an initial number I*(0) *of undetected infected cases, on the first day, from the internal evolution process we have I*(1) = *I*(0) + *Y*_1,0_ − *Y*_2,0_ − *Y*_5,0_, *then at the end of this day, we have I*^+^(1) = *I*(1)(1 + *p*), *where* 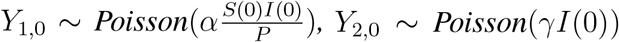 *and Y*_5,0_ ∼ *Poisson*(*βI*(0)). *Therefore, we have:* 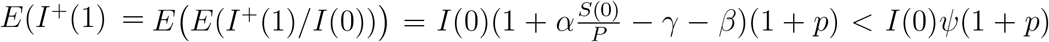. *Similarly, we have:* 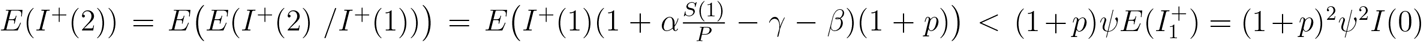. *Repeating this argument until reaching day T results in E*(*I*(*T*)^+^) < (1 + *p*)^*T*^ *ψ*^*T*^ *I*_0_. *For average control, therefore, we want to find a p such that:* (1 + *p*)*ψI*(0), (1 + *p*)^2^*ψ*^2^*I*(0), …, (1 + *p*)^*T*^ *ψ*^*T*^ *I*(0) < *c. By solving the above inequality, the p that satisfies the requirements is*

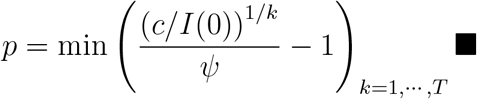
2. *Probability control. Here we have*

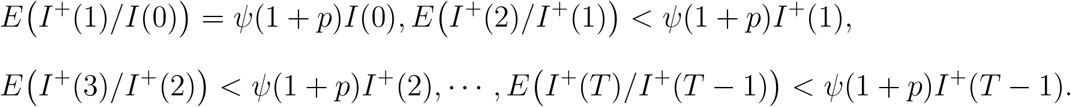

*Let us denote* 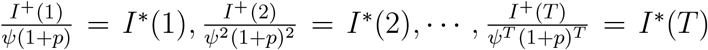. *The sequence I*^∗^(1), *I*^∗^(2), …, *I*^∗^(*T*) *forms a non-negative supermartingale sequence. Since*

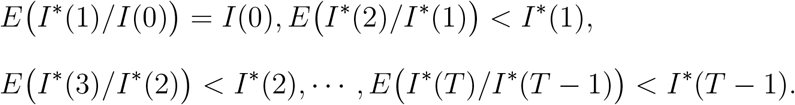

*Applying the maximal inequality for a non-negative supermartingale we have:* 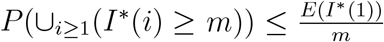, *for a given m >* 0. *This gives* 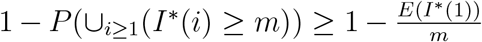. *In other words*, 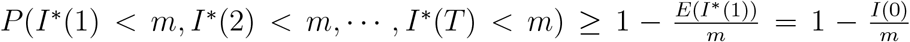. *If we want P (I*^∗^(1) < *m, I*^∗^(2) < *m*, …, *I*^∗^(*T*) < *m*) ≥ *p*_*c*_, *then the smallest value of m must satisfy the relation* 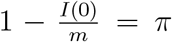. *We choose* 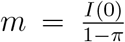. *We have: P (I*^∗^(1) < *m, I*^∗^(2) < *m*, …, *I*^∗^(*T*) < *m*) = *P I*^+^(1) < *mψ*(1 + *p*), *I*^+^(2) < *mψ*^2^(1 + *p*)^2^, …, *I*^+^(*T*) < *mψ*^*T*^ (1 + *p*)^*T*^. *So, if we want I*^+^(1), *I*^+^(2), …, *I*^+^(*T*) < *c, then we need to find a p such that mψ*(1 + *p*) < *c, mψ*^2^(1 + *p*) < *c*, …, *mψ*^*T*^ (1 + *p*)^*T*^ < *c. In other words, we need a p that satisfies:* 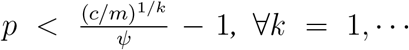, *T, or* 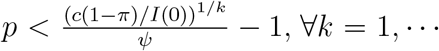, *T*. *In conclusion, for a probability control level π and a threshold c in the next T days, the required p is*

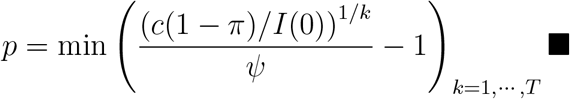

*Remark: The probability control policy is more conservative than the average control policy. Under the same threshold c, the difference between the two policies is the factor* (1 − *π) in the numerator of the probability control strategy. If we want the probability control to have at least 0*.*9 of the threshold c of average control, this factor becomes 0*.*1. As a result, the proportion of p in probability control is much smaller than the proportion of p in average control. If we want to use the probability control with a probability of at least 0*.*9, we need to set up the threshold c in probability control higher than the threshold c in average control to make sure that our p is non-negative*.

**Example 1**. Here we give one example of using the average control policy to regulate the travel. For simplicity, we consider a small world with only three countries, with the following initial states and true parameter values:

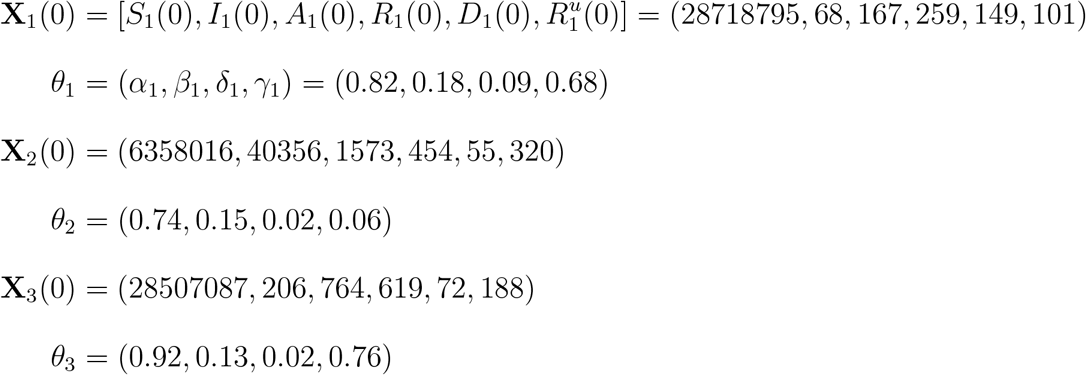

and we want to regulate the incoming travel in the first country (country 1). These choices of initial conditions and parameter values are based on our simulations where benefits of travel restriction can be seen clearly. We now need to find the regulation sequences {*p*_21_(*t*)}_*t*=1,…, 7_ and {*p*_31_(*t*)}_*t*=1,…, 7_ that can regulate airline travel from country 2 to country 1 and from country 3 country 1 such that for the next *T* = 7 days, the number of undetected infected cases in the arriving country will not exceed *c* = 70 cases on average per day. Applying Lemma 1a, we can find parameter *p* for country 1 as *p* = min ((*c/I*_0_)^1*/k*^*/ψ* – 1)_*k*=1,…, 7_ = 0.035.

So the sequence of the number of undetected infected imported cases that country 1 can accept each day as (((1 + *p*)*ψ*)^*i*^*I*(0)_*i*=1,…, 7_ = (2, 2, 2, 2, 2, 2, 2). Because country 1 has two “neighbors”, so country 1 can accept about 1 undetected infected from each neighbor each day during the regulation period.

The next step is to predict, for each day, the number of undetected infected travelers from countries 2 and 3 that can enter country 1 for the next 7 days when full travel is allowed. We get these numbers by simulating data given the true parameters under the fully open scenario. We first simulate 10000 stochastic realizations under this scenario and use the 0.975 percentile of the simulated sequence of undetected infected in countries 2 and 3 in the next 7 days as proxies for the number of undetected infected cases in these countries. Then we simulate a deterministic realization under the fully open scenario during the regulation period and use the values from the deterministic realization to calculate the percentage of undetected infected people in countries 2 and 3. Based on these percentages and the travel data, we estimate how many undetected infected individuals enter country 1 from country 2 and country 3 daily during the regulation period if full travel is allowed. The final step is obtaining the regulation sequence that country 1 can allow country 2 and country 3 to enter its border. The regulation sequence that country 1 can allow for country 2 to enter its border during the regulation period is obtained by dividing the number of daily undetected infected cases that country 1 can tolerate from country 2 by the daily estimated number of imported undetected infected cases from country 2 if full travel is allowed. Notice that if the daily proportion is greater or equal to 1, we set it to 1. Repeat the same procedure, we can also find the regulation sequence that country 1 can allow for country 3 to enter its border.

Following the above steps, we can find that in the next 7 days, the regulation sequence of proportions of people who can move from country 2 to country 1 is (0.103, 0.076, 0.051, 0.035, 0.022, 0.014, 0.009), and the sequence of proportions of people who can move from country 3 to country 1 is (1, 1, 1, 1, 1, 1, 1). Compared to the fully open scenario, using the average control approach with the threshold of 70 cases during the 7-day regulation period, about 6.03% of travelers from country 2 are allowed enter the country 1, and all travelers from country 3 are allowed to enter country 1. Overall, the volume of inbound travelers in country 1 is about 88.64% of the normal levels.

In practice, the value of the transmission rate *α* varies over time, and we therefore provide an additional lemma that generalizes Lemma 1 to address the aspect of varying *α*.

##### Lemma 2.

*Under the assumptions of our model, for a given country with population size P, initial status* **X**(0) = [*S*(0), *I*(0), *A*(0), *R*(0), *D*(0), *R*^*u*^(0)] *and the parameter of the statistical model for this country over the time period from* 0 = *T* (0) *to T* = *T (m) is θ* = (*α, β, δ, γ*), *where α* = *α*_1_ℐ_0,*T* (1)_(*t*) + *α*_2_ℐ_*T*(1),*T* (2)_(*t*) + …+ *α*_*m*_ℐ_*T*(*m*−1),*T*(*m*)_(*t*). *Let us denote* 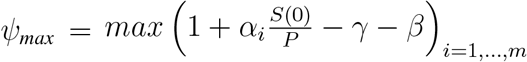.

1. *The average control requirement is satisfied if*

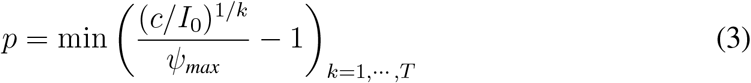
2. *The probability control requirement is satisfied if*

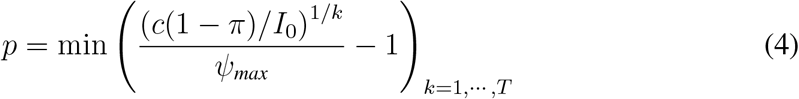

*Proof:*

1. *Average control. Follow the same argument as in the proof of Lemma 1, we have* 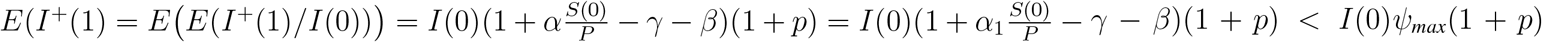. *Repeating the argument until we reach day T yields* : 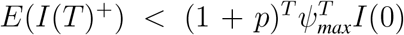. *Therefore, we want to find a p such that* 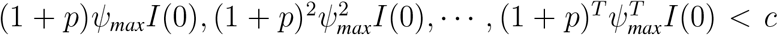. *Hence the value p that satisfies the requirements for average control is*

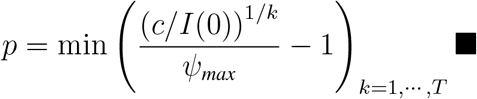
2. *Probability control. Here we have*

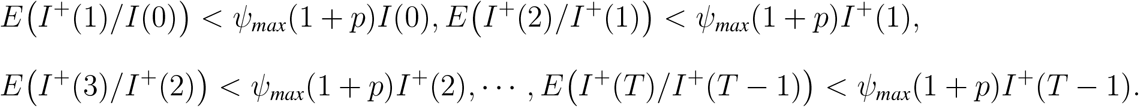

*Let us denote* 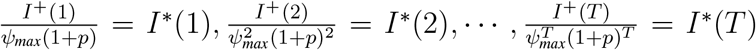. *Similar to Lemma 1, the sequence of I*^∗^(1), *I*^∗^(2), …, *I*^∗^(*T*) *forms a non-negative supermartingale sequence. Hence, following the same arguments as in Lemma 1, we have for a probability control level π and a threshold c in the next T days, the required p is*

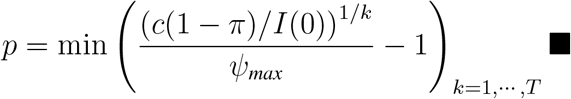

#### 1.4 Choosing distance and summary statistics in Approximation Bayesian Computational

There are many variants of ABC, but they are all based on a comparison of observed and simulated data, which in most cases requires specification of data summary statistics, a distance measure, and a scalar distance threshold *ϵ*. The most basic ABC algorithm, the so-called accept-reject method, starts by simulating a parameter value from a prior distribution and then uses the model, given this parameter value, to generate one realization of data. If the distance between the summary statistics for the observed data and the summary statistics for the simulated data is less than or equal to *ϵ*, the sampled parameter value is retained; otherwise, it is discarded. The collection of accepted parameter values constitutes a sample from an approximation of the posterior distribution. The approximation generally improves with smaller values of *ϵ*, but at the same time it becomes more computationally expensive to obtain acceptances.

This basic ABC algorithm is computationally inefficient when working with a small threshold *ϵ* as a vast majority of sampled parameter values are rejected. To address this inefficiency, some sequential variants of ABC have been proposed, such as the ABC Markov chain Monte Carlo algorithm (ABC-MCMC) by Marjoram, et al. (2003) and the ABC Sequential Monte Carlo algorithm (ABC-SMC) by Sisson et al. (2007), Toni, et al. (2009), and Drovandi and Pettitt (2011) (*3–6*). In this paper, we use the variant from Drovandi and Pettitt (2011) (*6*), called replenishment ABC (RABC). For its implementation, we use the R package protoABC from Ebert (2020) (*7*). This package is very flexible as the users can employ any priors, generative models, and distance functions.

A commonly used distance is the Euclidean distance due to its simple form. In our problem setting, this Euclidean distance *L (S* Data^(*i*)^), *S*(Data) can be written as:

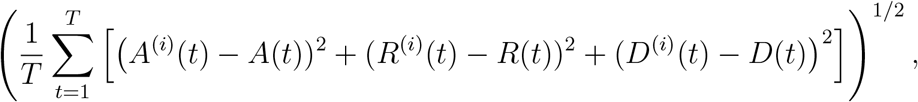

where *A*^(*i*)^(*t*), *R*^(*i*)^(*t*), *D*^(*i*)^(*t*) and *A*(*t*), *R*(*t*), *D*(*t*) are active confirmed, accumulated recovered confirmed, and accumulated death confirmed cases on day *t* of the simulated data and the real (empirical) data, respectively. However, simply using the Euclidean distance may not be the best choice since it does not account for the scale of different quantities, and may need to be standardized (see for example Beaumont et al. (2002), Csilléry et al. (2012), or Prangle (2017) (*8–10*)). This is why we also consider the following weighted Euclidean distance, with weights given by the inverse variances:

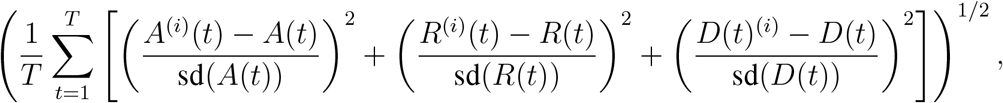

where {sd(*A*(*t*)), sd(*R*(*t*)), sd(*D*(*t*))}_*t*=1,…, *T*_ are the prior predictive standard deviations of *A*(*t*), *R*(*t*), *D*(*t*) at each time step. These standardization values are obtained by first generating *N (N* being large) parameters values *θ* = (*α, β, δ, γ*) from the prior *π*(*θ*), and then generating one realization of simulated data for each of them. The standard deviation at each time step is then calculated based on these *N* simulated data, giving {sd (*A*(*t*)), sd (*R*(*t*)), sd (*D*(*t*))}_*t*=1,…, *T*_. In our simulation study, we choose *N* = 5000.

To improve the predictive quality of ABC algorithms, we can also use additional information from parameter estimates to make new summary statistics. Under our model assumptions, we can learn additional knowledge about how parameters can be estimated. We are thus going to include as summary statistics estimates of our epidemiological parameters. For simplicity, we first limit our discussion to the local model, where each country is considered separately. The choice of the distance for the global model will be discussed in Section 1.5.

Under our model assumptions, we have: *R*(*t*) = *R*(*t* − 1) + Poisson (*βA*(*t* − 1)), ∀*t* = 1, …, *T*. So for a given *A*(*t* − 1), we have: *E (R*(*t*) − *R*(*t* − 1)) = *βA*(*t* − 1). This yields 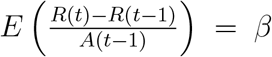. So for a given sequence of known {*A*(*t*)}_*t*=1,…, *T*_, the sequence of independent variables 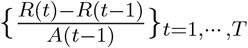 has *β* as common mean value. Therefore, we can use its median value to estimate *β*. If our algorithm generated a reasonable *θ*^(*i*)^, then the data generated by *θ*^(*i*)^ should also gives us a sequence 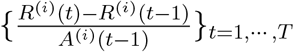 with median value close to the corresponding median value of 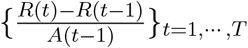. Therefore under our model assumptions, adding the term 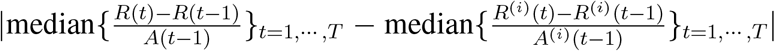 when calculating *L S*(Data^(*i*)^), *S*(Data) would help to improve the estimation of *β*. Similarly, the median of the sequence 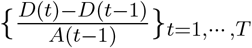 can be used to estimate the death rate *δ*, and adding the term 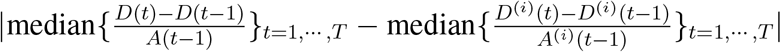 would help to improve the estimation of *δ*.

We now try to learn the transmission rate *α* under our model assumptions. We have *S*(*t*) = *S*(*t* − 1) + Poisson 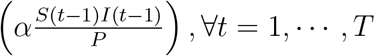, where *P* is the total population size of the country. So for a given *S*(*t* − 1), *I*(*t* − 1), we have 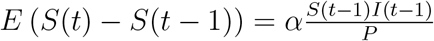. This yields 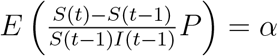. Unfortunately, *S*(*t* − 1) and *I*(*t*−1) are hidden states and not available in our data. To use the above strategy to improve the estimation of *α*, we need to reconstruct these hidden states based on the available data {(*A*(*t*), *R*(*t*), *D*(*t*))}_*t*=1,…, *T*_. Because our model is stochastic, all values would change each time we rerun the model. However, based on the available data {(*A*(*t*), *R*(*t*), *D*(*t*))}_*t*=1,…, *T*_ we can reconstruct the mean realization that adopts these three states. Let us denote *U (t*) the total number of confirmed cases at time *t*, Δ*U (t* − 1) the number of new confirmed cases at time *t* as, and Δ*R*^*u*^(*t* − 1) the number of new undocumented recover cases at day *t*. Note that *U (t*) = *A*(*t*) + *R*(*t*) + *D*(*t*), Δ*U (t* − 1) = *U (t*) − *U (t* − 1), and Δ*R*^*u*^(*t* − 1) = *R*^*u*^(*t*) − *R*^*u*^(*t* − 1).

From the local model we have:

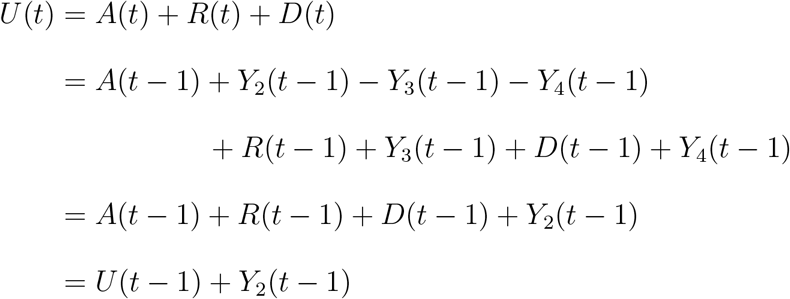

So *Y*_2_(*t* − 1) = *U (t*) − *U (t* − 1) = Δ*U (t* − 1). Moreover, since *Y*_2_(*t* − 1) ∼ Poisson (*γI*(*t* − 1)), we have

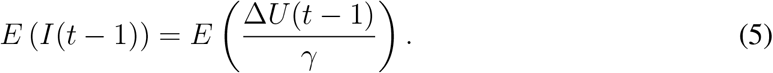

Equation (5) tells us that with the observed data {*A*(*t*), *R*(*t*), *D*(*t*)}_*t*=1,…, *T*_, if the identification rate *γ* is known, the average realization *I*(*t*) can be reconstructed up to time *T* − 1.

Similarly, since *R*^*u*^(*t*) = *R*^*u*^(*t* − 1) + *Y*_5_(*t* − 1), where *Y*_5_(*t* − 1) ∼ Poisson (*βI*(*t* − 1)), we have

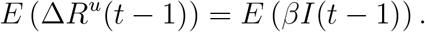

Using (5), we obtain

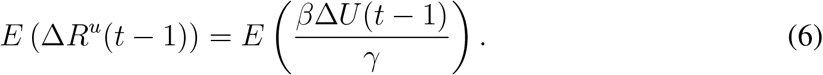

Moreover, using 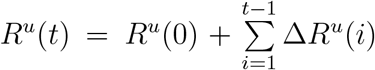 and (6), the average value of *R*^*u*^(*t*) can be reconstructed as

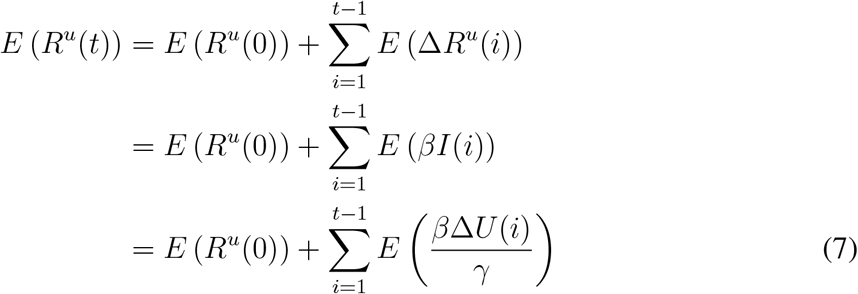

Equations (5) and (7) tell us that based on the available data {(*A*(*t*), *R*(*t*), *D*(*t*)}_*t*=1,…, *T*_ if the identification rate *γ* and the recovered rate *β* are available to us, then we can reconstruct the average realization of *I*(*t*) and *R*^*u*^(*t*), ∀*t* = 1, …, *T* − 1. As a result the average realization category at time *t* can also be reconstruct as 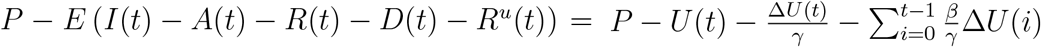, where *P* is the country population.

Overall, based on the observed data {(*A*(*t*), *R*(*t*), *D*(*t*))}_*t*=1,…, *T*_, suppose that the identification rate *γ* and the recover rate *β* are available to us. The average realization that adopts {(*A*(*t*), *R*(*t*), *D*(*t*))}_*t*=1,…, *T*_ can be reconstructed up to time *T* − 1 as

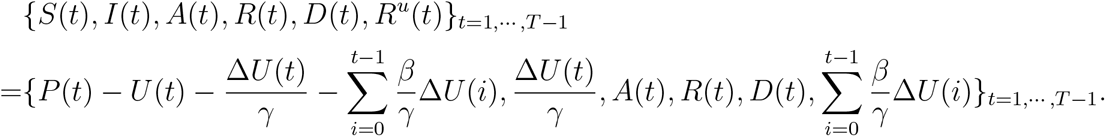

Therefore *α* can be estimated as the median of the sequence 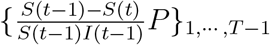. Unfortunately, *β* and *γ* are not known in advance and need to be estimated.

We therefore use the testing argument to recover the average realization and estimate *α*. This argument is based on the following observation. In step 1 of the ABC algorithm, a parameter value *θ*^(*i*)^ = *α*^(*i*)^, *β*^(*i*)^, *δ*^(*i*)^, *γ*^(*i*)^ is generated and available to us. If *γ*^(*i*)^, *β*^(*i*)^ are correctly specified as *γ, β* of the underlying true parameter *θ* = (*α, β, δ, γ*). We would expect that the median value of the sequence 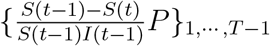 constructed using *γ*^(*i*)^, *β*^(*i*)^ and the available data {(*A*(*t*), *R*(*t*), *D*(*t*))}_*t*=1,…, *T*_, should give us a good estimator for the underlying true *α* value. A similar statement holds for the median value of the sequence 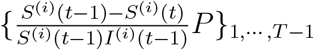 that was constructed by *γ*^(*i*)^, *β*^(*i*)^ and the available data {(*A*^(*i*)^(*t*), *R*^(*i*)^(*t*), *D*^(*i*)^(*t*)}_*t*=1,…, *T*_ should also give us a good estimator for the underlying true *α* value. Therefore the distance 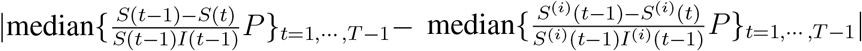 in *L (S*(Data^(*i*)^),*S*(Data)) should be close to 0. On the other hand, if the generated parameters *γ*^(*i*)^, *β*^(*i*)^ are far away from the underlying true parameters, then 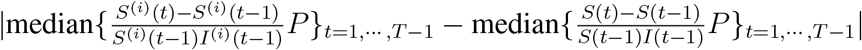 should not be close to 0.

Based on this observation, we then add the term

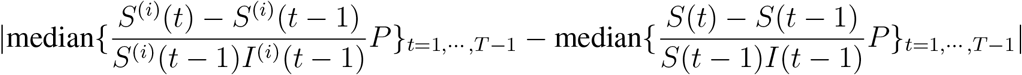

in *L(S*(Data^(*i*)^), *S*(Data)) to improve the estimation for *α*.

Finally, our proposed distance for the model is designed as follows:

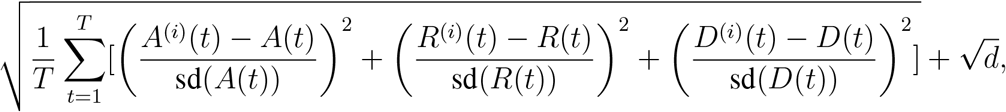

Where

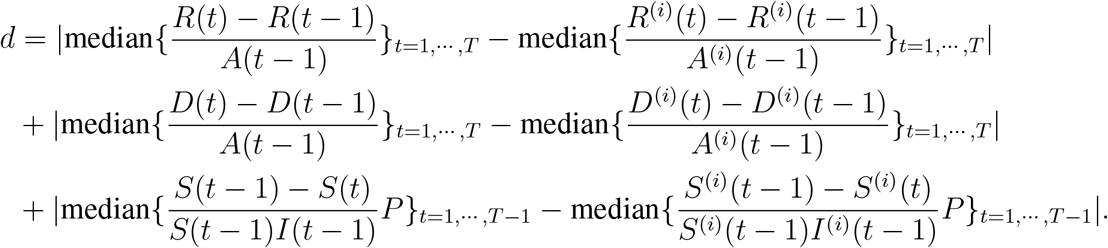

#### 1.5 Marginal approach to parameter estimation

In the following, we will discuss how to use ABC to estimate parameters in each country for our global model. The challenge of using ABC to estimate the global model parameters is that many parameters need to be estimated. Therefore directly using ABC to estimate all the parameters for all countries at once may result in very unstable parameter estimations and will be extremely computationally intensive. We propose a marginal estimating approach to estimate each country parameter for the global model separately while still taking into account the travel data.

For simplicity, let us first consider a given country *m* with the represented parameter *θ*_*m*_ = (*α*_*m*_, *β*_*m*_, *δ*_*m*_, *γ*_*m*_). Let us denote *A*_*k*_(*t*), *R*_*k*_(*t*), *D*_*k*_(*t*) as the number of active confirm cases, accumulated recover confirmed, and accumulated death confirmed at country *k* on day *t*, respectively. We denote *T (t*) = [*T*_*ij*_(*t*)]_*i,j*=1,…, *n*_ is the global traveling matrix at day *t*, where *T*_*ij*_(*t*) is the number of travelers from country *i* to country *j* at day *t*. Notice that *T*_*ij*_(*t*) = 0 if *i* = *j*, ∀*t* ∈ 1, …, *T*. With the available data from the global model {*A*_*k*_(*t*), *R*_*k*_(*t*), *D*_*k*_(*t*)}_*k*=1,…, *n*;*t*=1,…, *T*_ and the travel data {*T (t*)}_*t*=1,…, *T*_, we need to estimate *θ*_*i*_.

Before introducing our estimation procedure, we rewrite how our global model evolved for the country *m* at day *t* and (*t* + 1): the country *m* at day *t* and (*t* + 1):

Step 1a *The internal pandemic evolves at day t:* The internal pandemic situation in the country evolves from 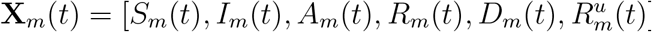, where *S*_*m*_(*t*) = *S*_*m*_(*t* −1) −*Y*_1,*m*_(*t* −1), *I*_*m*_(*t*) = *I*_*m*_(*t* −1) + *Y*_1,*m*_(*t* −1) −*Y*_2,*m*_(*t* −1) −*Y*_5,*m*_(*t* −1), *A*_*m*_(*t*) = *A*_*m*_(*t* − 1) + *Y*_2,*m*_(*t* − 1) − *Y*_3,*m*_(*t* − 1) − *Y*_4,*m*_(*t* − 1), *R*_*m*_(*t*) = *R*_*m*_(*t* − 1) + *Y*_3,*m*_(*t* − 1), *D*_*m*_(*t*) = *D*_*m*_(*t* − 1) + *Y*_4,*m*_(*t* − 1), 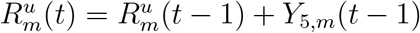. And *Y*_*j,m*_(*t* − 1), *j* = 1, …, 5 are Poisson distributed with rates (*h*_*j,i*_(**X**_*m*_(*t* − 1))), respectively, as 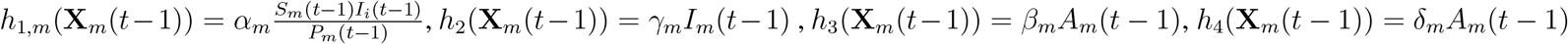, and *h*_5_(**X**_*m*_(*t* − 1)) = *β*_*m*_*I*_*m*_(*t* − 1).

Step 1b *The external pandemic added at day t:* From the travel data, **X**_*m*_(*t*) is updated to 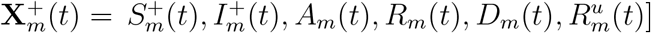, where 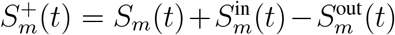, and 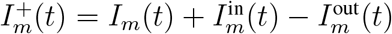.

Step 2a *The internal pandemic evolves at day (t* + 1): The internal pandemic situation in the country evolves from 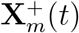 to 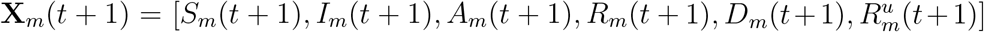 as 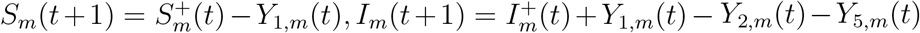, *A*_*m*_ (*t*+1) = *A*_*m*_(*t*) + *Y*_2,*m*_(*t*)−*Y*_*3,m*_(*t*)− *Y*_4,*m*_(*t*),*R*_*m*_(*t*+1) = *R*_*m*_(*t*)+ *Y*_3,*m*_(*t*),*D*_*m*_(*t* + 1) = *D*_*m*_(*t*) + *Y*_4,*m*_(*t*), 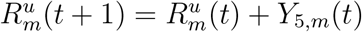. And *Y*_*j,m*_(*t*), *j* = 1, …, 5 are Poisson distributed with rates *h*_*j,m*_(**X**_*m*_(*t*)), respectively, as 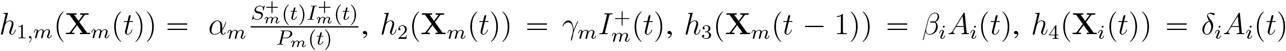, and 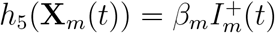.

Step 2b *The external pandemic added at day (t* + 1): From the travel data, **X**_*m*_(*t* + 1) is updated to 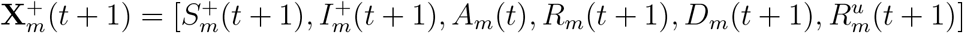, where 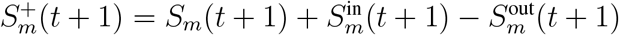, and 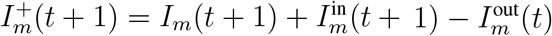.

As shown in the model’s evolving process, step 1b and step 2b make the global model behave differently from the local model. Therefore, if we can estimate quantities 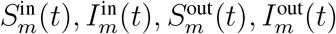 for each day *t*, then we can use the marginal approach to estimate each country’s parameters separately. The two quantities 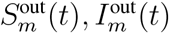 come from inside the considered country *m*. Therefore the amount can be calculated during the data generation process of the ABC algorithm. Our main task now is estimating 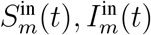.

We have 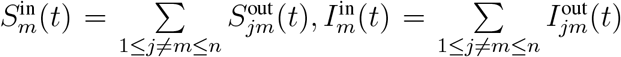, where 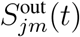 and 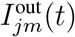 are the number of susceptible people and undocumented infected people move from country *j* to country *m* at day *t*, respectively. It should be noticed that under our model assumptions the summation of 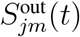 and 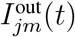 gives us the total number of people traveling from country *j* to country *m* at day *t*, i.e. 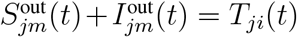. So if we can estimate 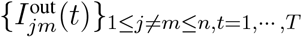 then with the travel data, we can estimate 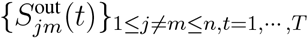. As a result we can then estimate 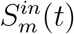, and 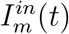.

We discuss the procedure for estimating 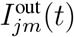. To estimate 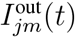 we need to estimate the pandemic situation in country *j* at day *t*, i.e. we need to estimate:

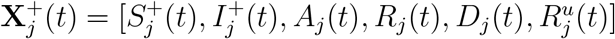. Then based on these compartments and the travel data *T*_*jm*_ (*t*), we can estimate 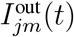 as 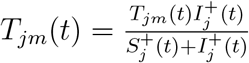.

From the global model we have:

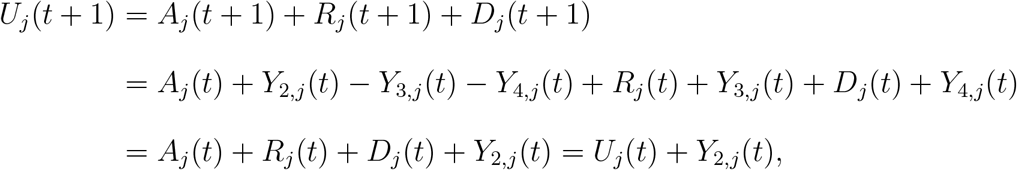

so *Y*_2,*j*_(*t*) = *U*_*j*_(*t* + 1) − *U*_*j*_(*t*) = Δ *U*_*j*_(*t*).

Since 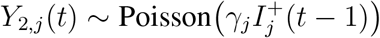, therefore, we have

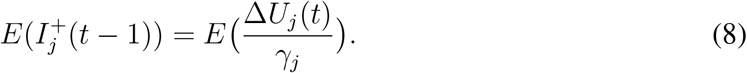

Similarly we also have

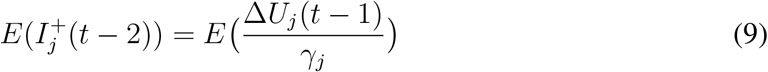

From (8) and (9), we have

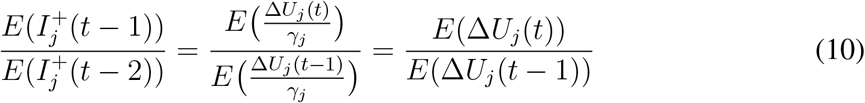

Applying (10) for *t* = 1, …, *T* − 1, we have the sequence of relationships: 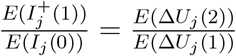, 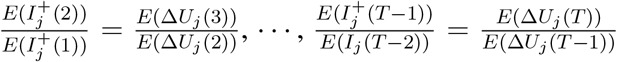. Therefore, based on the available data at country *j* as {*A*_*j*_(*t*), *R*_*j*_(*t*), *D*_*j*_(*t*)}_*t*=1,*…,T*_, we can approximate the average realization of the sequence of undetected infected people in country *j* up to time (*T* − 1) by *I*_*j*_(0), 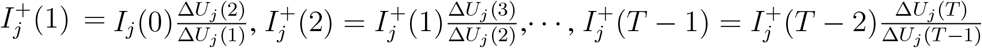.

In addition, we also have 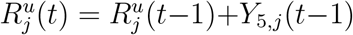, where 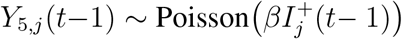. Therefore,

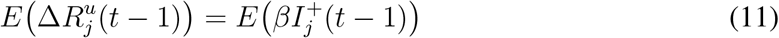

We have *R*_*j*_(*t*) = *R*_*j*_(*t* − 1) + *Y*_3,*j*_(*t* − 1), where *Y*_3,*j*_(*t* − 1) ∼ Poisson *βA*_*j*_(*t* − 1), ∀*t* = 1, …, *T*. Therefore the median value of the sequence 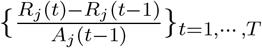 can be used to approximate *β*. We denote this median value as 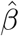.

The fact 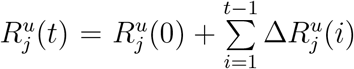 and (11) tells us that the average value of *R*^*u*^ (*t*)at country *j* can be reconstructed as

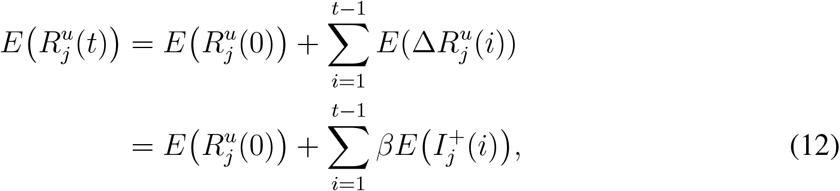

where the sequence 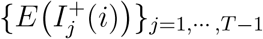 and *β* are estimated as above.

So the average realization of the pandemic at country *j* which adopts {(*A*_*j*_(*t*), *R*_*j*_(*t*), *D*_*j*_(*t*)}_*t*=1,*…,T*_ as its active confirmed, recover confirmed and death confirmed can be reconstructed up to time *T* − 1 as 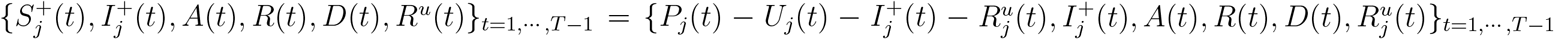.

This procedure of estimating the average realization of a given country *j* based on the available data {(*A*_*j*_(*t*), *R*_*j*_(*t*), *D*_*j*_(*t*))}_*t*=1,*…,T*_ is completed. As a result, this gives us the estimation of 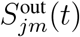 and 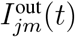. This means we can estimate 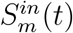, and 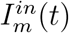. Therefore, the underlying true parameter *θ*_*m*_ in a given country *m* can be approximated marginally by using the above estimating procedure.

We now discuss the proposed distance when estimating *θ*_*m*_ in a given country *m* by ABC marginally. Following the same argument as in Section 1.4, instead of using the commonly used Euclidean distance to estimate *θ*_*m*_ we first need to standardize each sequence, then we try to learn each parameter under our model assumptions. From the available data {(*A*_*m*_(*t*), *R*_*m*_(*t*), *D*_*m*_(*t*))}_*t*=1,*…,T*_ of country *m*, follow the same argument as in Section 1.4, we can add the term 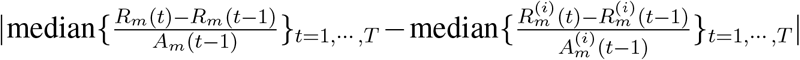 to improve the estimation for the recover rate *β*_*m*_, and adding the term 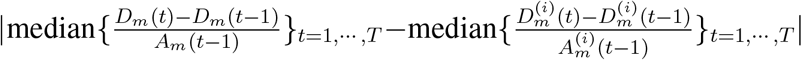 to improve the estimation for the death rate *δ*_*m*_. We discuss the transmission rate *α*_*m*_, at a time step *t* + 1, we have: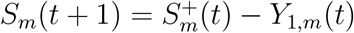 where 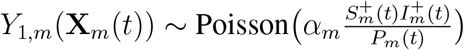. Therefore 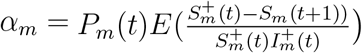. Notice that 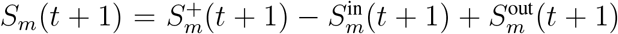. The hidden states 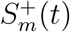 and 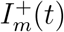 can be reconstructed as above or by using the testing argument as above. We discuss the approach by using the testing argument.

From (8) we have: 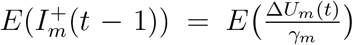. Therefore the average realization of 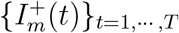 can be reconstructed as 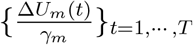. Using (12), we have 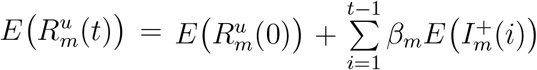. Therefore the average realization of 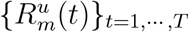 can be reconstructed as 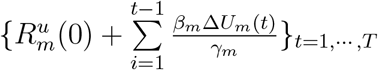

The average realization of the pandemic at country *m* which adopts {(*A*_*m*_(*t*), *R*_*m*_(*t*), *D*_*m*_(*t*)}_*t*=1,*…, T*_ as its active confirmed, recover confirmed and death confirmed can be reconstructed up to time *T* − 1 as 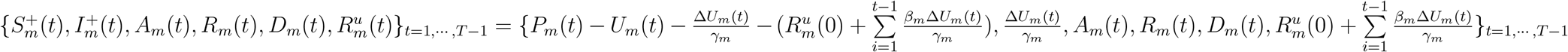.

Similarly as above, in step 1 of the ABC algorithm, the parameter 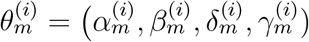 is generated and available to us. If 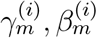 are correctly specified as *γ*_*m*_, *β*_*m*_ of the underlying true parameter *θ*_*m*_ = (*α*_*m*_, *β*_*m*_, *δ*_*m*_, *γ*_*m*_), we would expect the distance 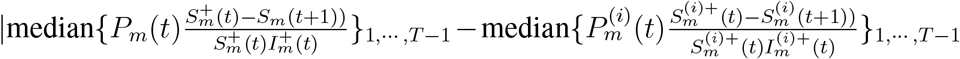 to be close to 0. Where values of the sequence 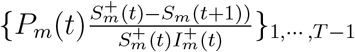 are constructed based on 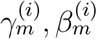 and the available data {(*A*_*m*_(*t*), *R*_*m*_(*t*), *D*_*m*_(*t*))}_*t*=1,*…,T*_, and values of the sequence 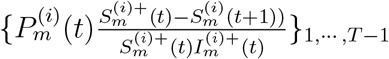 are constructed based on 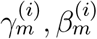 and the simulated data 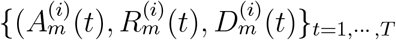. So adding the term: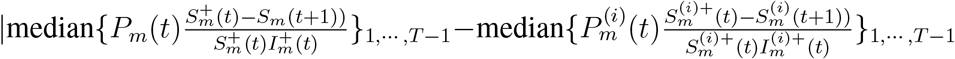 would help to improve the estimation for *α* _*m*_. Finally, the proposed global distance in the calibrating step of the ABC algorithm is designed as follow:

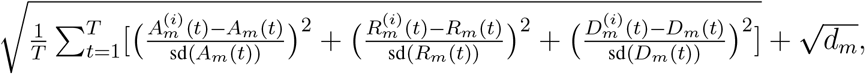

Where

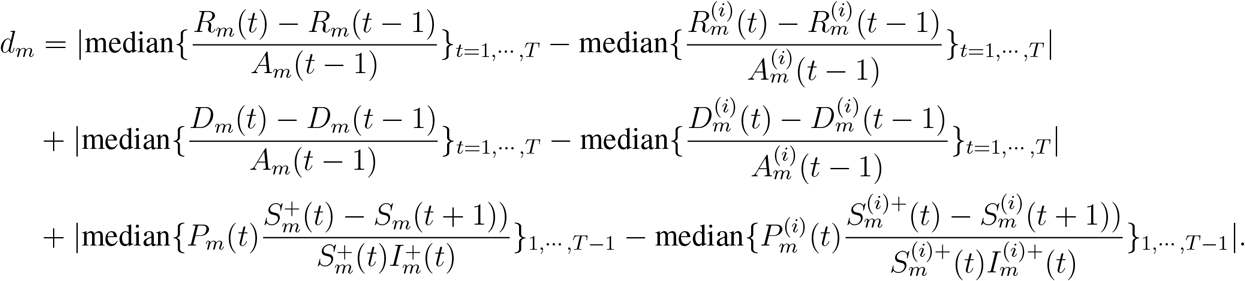

### 2 Simulation studies

#### 2.1 Simulation 1: Performance of different RABC distance metrics

For our first simulation study, we limit our model to the analysis of only one country, i.e., we only use the internal model. We here demonstrate the impact of the choice of the distance in ABC algorithms and which one to choose in our epidemiological framework.

We simulate *N* = 200 sets of parameters and data, in an ABC fashion, by first simulating a parameter value from the prior and using it to generate data according to the model. We treat these *N* simulations as our test data set to assess how accurately the true parameters are recovered by ABC using various distance functions. The simulation proceeds as follows.

*Step 1. Generating data and parameters*: For *i* ∈ {1, …, *N*} (N large), we generate the parameter *θ*^(*i*)^ = (*α* ^(*i*)^, *β*^(*i*)^, *δ*^(*i*)^, *γ*^(*i*)^) from uniform priors *α* ^(*i*)^ ∼ *U* (0, 2), *β*^(*i*)^ ∼ *U* (0, 1), *δ*^(*i*)^ ∼ *U* (0, 1), and *γ*^(*i*)^ ∼ *U* (0, 1). Based on the parameters and the stochastic model, we generate a data set Data^(*i*)^ corresponding to *θ*^(*i*)^. If the generated data set Data^(*i*)^ satisfies certain conditions making it sufficiently real-world like, such as having the number of confirmed accumulated deaths greater than 1% and lower than 30% of total confirmed cases and having the number of accumulated recovered cases at least twice the number of accumulated deaths, then we keep *θ*^(*i*)^ as a true parameter value to be estimates and treat the generated data 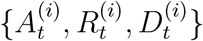 as real observed data. We repeat the process until we obtain 200 underlying true parameter values *θ*^(*i*)^ and the corresponding 200 datasets 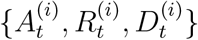. For simplicity, we fix the initial condition of the six compartments in the model as 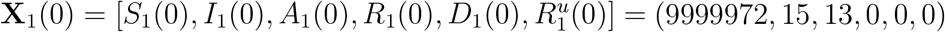 and set the simulation time period *T* = 84 days for all *i*.

*Step 2. Estimating parameters*: For each iteration *i, i* … {1, …, 200}, based on the sequence of 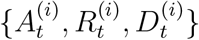, we use RABC with different distance metrics to estimate the underlying true parameter value *θ*^(*i*)^. In this estimation step, we choose the acceptance rate 0.01 and sample 1000 particles to form the posterior. From the posterior distribution for each *i*, we calculate the median values of each parameter: 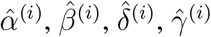. Then 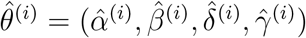 is used as the best candidate for estimating the underlying true *θ*^(*i*)^.

*Step 3. Evaluating parameter estimates*: For each iteration *i, i* … {1, …, 200}, we evaluate estimation accuracy in terms of the absolute bias, absolute relative bias, interquartile range, and coverage rate of the interquartile for each parameter *α* ^(*i*)^, *β*^(*i*)^, *δ*^(*i*)^, *γ*^(*i*)^ and its average. These accuracy measurements are defined as follows. For a given parameter *α* ^(*i*)^ the absolute bias is defined as 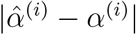, and the absolute relative bias is defined as 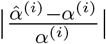. Similarly for *β*^(*i*)^, *γ*^(*i*)^, and *δ*^(*i*)^. Average absolute bias for all parameters is defined as 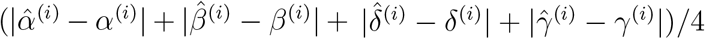 and average absolute relative bias for all parameters is defined as 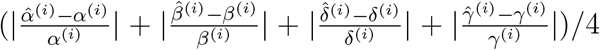. For each parameter, we also calculate the interquartile range (IQR) of the posterior, denoted IQR^(*i*)^, which is the difference between the third and the first quartile of the resulting ABC posterior distribution. Furthermore, we check whether the IQR of the posterior covers the underlying true parameter value, which we use to calculate the coverage rate CR^(*i*)^. Then the average of the IQR for all four parameters and the average of the coverage rate is calculated to characterize the overall performance of the two distance metrics. Finally, the average over the 200 iterations of these accuracy metrics is calculated, which we use as our overall accuracy metrics for comparing the performance of the two RABC distance metrics.

Table S1 summarizes the different prediction accuracy measures for the two distances. This table shows that the distance we proposed increases the estimation accuracy in terms of relative bias. The two types of bias are much smaller compared to using Euclidean distance. We also observe that the IQR for the proposed distance is higher than the IQR for the Euclidean distance, but the proposed distance also yields more narrow IQR. This means that our proposed distance metric more frequently correctly bounds the true parameter values. The IQR is about 2.5 times smaller when using the proposed distance metric instead of Euclidean distance. Figure S1 shows boxplots of the IQR for the two distance metrics.

**Table S1:** Accuracy of Euclidean distance and our proposed distance when using RABC to estimate the four parameters of the local model.

| Accuracy | Distances | Average | alpha | beta | delta | gamma |
| --- | --- | --- | --- | --- | --- | --- |
| Absolute bias | Euclidean | 0.071 | 0.125 | 0.046 | 0.012 | 0.101 |
|  | Proposed | 0.028 | 0.054 | 0.004 | 0.002 | 0.052 |
| Absolute relative bias | Euclidean | 0.148 | 0.107 | 0.089 | 0.093 | 0.303 |
|  | Proposed | 0.077 | 0.039 | 0.007 | 0.022 | 0.241 |
| IQR | Euclidean | 0.153 | 0.264 | 0.094 | 0.031 | 0.222 |
|  | Proposed | 0.060 | 0.107 | 0.014 | 0.012 | 0.109 |
| IQ coverage | Euclidean | 0.677 | 0.620 | 0.675 | 0.770 | 0.645 |
|  | Proposed | 0.777 | 0.635 | 0.880 | 0.960 | 0.635 |

**Figure S1:**
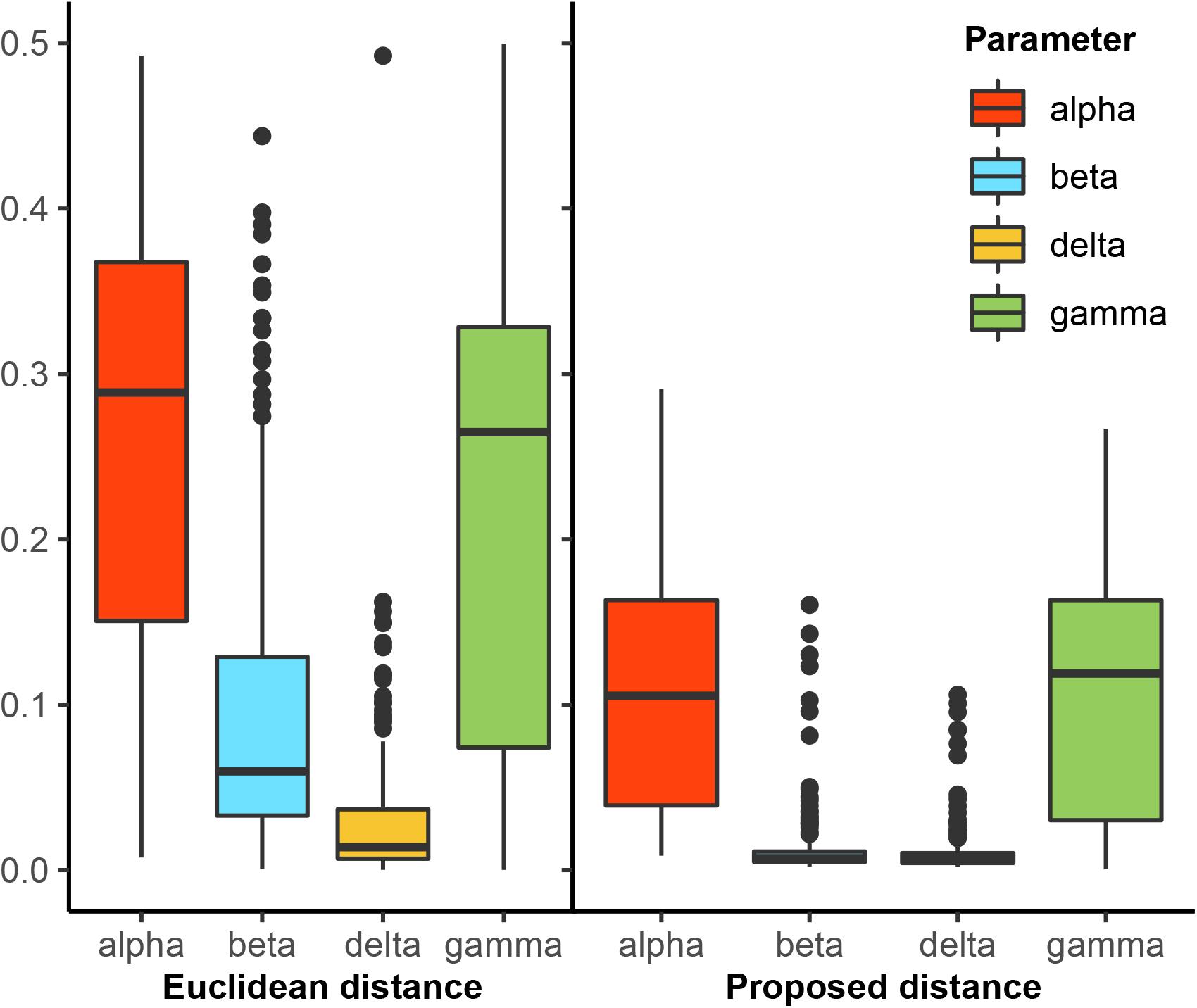
Interquartile range of the posterior for parameters estimation of the proposed distance and the Euclidean distance for the local model of one country.

#### 2.2 Simulation 2: Performance of local and global estimation

In this simulation study, we investigate the accuracy of three different estimation procedures for the global travel model consisting of three countries. (We limit this investigation to three countries for simplicity.) The first procedure uses a local approach with the Euclidean distance to estimate each country’s parameters independently and ignores the travel between the countries. We call this estimation procedure Euclidean local, and we use it as a benchmark to be compared with the other two approaches. Then we consider a global estimation procedure as discussed in Section 1.5 to estimate each country’s parameters. Here we use two distance metrics, Euclidean distance and the distance proposed in Section 1.5. We call these estimation procedures Euclidean global and Proposed global, respectively. The simulation is set up as follows.

*Step 1. Generating data and parameters*: For *i* ∈ {1, …, *N*} (N large), we generate the parameter 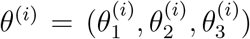, where for each *j* in 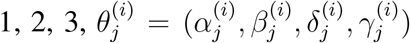 from uniform priors as 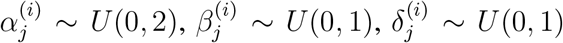, and 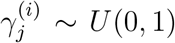. Based on the parameters and the stochastic model, we generate a data set Data^(*i*)^ corresponding to *θ*^(*i*)^. If the generated data set Data^(*i*)^ satisfies the conditions described above for Simulation 1, we retain *θ*^(*i*)^ and treat it as the underlying true parameter value; we also retain the data 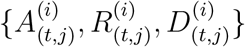, for *j* = 1, 2, 3, and treat them as the observed data from these three countries. We repeat the procedure until we have 500 parameter values and their corresponding data sets 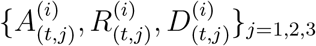.

For simplicity, we fix the initial condition of the six compartments in the model as

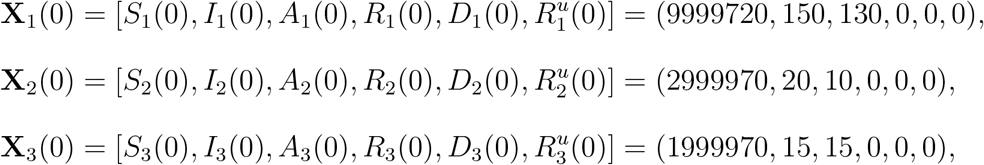

and set the simulation period *T* = 84 days for all *i*. Each day, the number of outbound travelers from country *j, j* = 1, 2, 3, is drawn from a normal distribution with mean *µ*_*j*_ = *P*_*j*_ ∗ 0.0003 and standard deviation sd_*j*_ = 0.05 ∗ *µ*_*j*_, where *P*_*j*_ is the size of the population of country *j*. Those outbound travelers will enter one of the neighboring countries with proportions that are proportional to the sizes (populations) of the target countries. For example, if there are *n*_1_ people leaving country 1, the number of them entering country 2 is 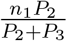 and the number of them entering country 3 is 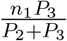.

*Step 2. Estimating parameters*: For each iteration *i, i* ∈ {1, …, 500}, based on the sequence of 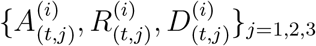, we first naively use RABC with the local estimation approach and Euclidean distance to estimate *θ*^(*i*)^. Then we use RABC with the global estimation approach with the two distance metrics to estimate the underlying true parameters *θ*^(*i*)^. Then 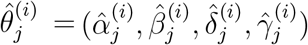 is obtained as the median of the RABC posterior samples and is used to estimate the underlying true 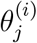, for *j* = 1, 2, 3.

*Step 3. Evaluating parameter estimates*: For each iteration *i, i* ∈ {1, …, 500}, for each country, we evaluate the accuracy of our parameter estimates based on the absolute bias, absolute relative bias, interquartile range (IQR), and coverage rate of IQR for each parameter *α*^(*i*)^, *β*^(*i*)^, *δ*^(*i*)^, *γ*^(*i*)^ and its average as in simulation 1. The final accuracy measurements are calculated by averaging the accuracy measurements across all three countries. When averaging accuracy measures over multiple countries, we consider two weighted averages, one having equal weights for all countries regardless of their population sizes and the other weighted based on relative population sizes. In the latter, the weights are 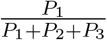 for country 1, 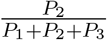 for country 2, and 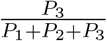 for country 3.

Tables S2 and S3 show the overall accuracy of different estimation procedures using equal weights for each country (Table S2) and using population-based weights for each country (Table S3). The two tables convey the same message: using a local approach to estimate the parameters in the travel model is not appropriate. As shown in these tables, the Euclidean local estimation procedure yields the highest bias, largest interquartile range, largest 95 % percentile range, and lowest coverage. The performance is better for the Euclidean global procedure. As expected, the proposed distance, which takes into account the travel model, performs best of the three.

**Table S2:** Accuracy of Euclidean distance and our proposed distance when using RABC to estimate the four parameters of the global model. For simplicity, we consider a small world of just three countries with different population sizes; here each country has the same weight when computing the overall accuracy.

| Accuracy | Estimation procedure | Average | alpha | beta | delta | gamma |
| --- | --- | --- | --- | --- | --- | --- |
| Absolute bias | Euclidean local | 0.496 | 1.328 | 0.108 | 0.058 | 0.490 |
|  | Euclidean global | 0.236 | 0.448 | 0.070 | 0.030 | 0.397 |
|  | Proposed global | 0.205 | 0.497 | 0.023 | 0.054 | 0.248 |
| Absolute relative bias | Euclidean local | 0.928 | 1.611 | 0.176 | 0.591 | 1.332 |
|  | Euclidean global | 0.502 | 0.534 | 0.109 | 0.280 | 1.085 |
|  | Proposed global | 0.494 | 0.629 | 0.034 | 0.545 | 0.769 |
| IQR range | Euclidean local | 0.171 | 0.263 | 0.179 | 0.060 | 0.180 |
|  | Euclidean global | 0.149 | 0.218 | 0.129 | 0.040 | 0.210 |
|  | Proposed global | 0.091 | 0.135 | 0.041 | 0.038 | 0.151 |
| IQ coverage | Euclidean local | 0.528 | 0.463 | 0.577 | 0.690 | 0.381 |
|  | Euclidean global | 0.610 | 0.539 | 0.649 | 0.714 | 0.540 |
|  | Proposed global | 0.631 | 0.543 | 0.647 | 0.761 | 0.574 |
| 95% range | Euclidean local | 0.410 | 0.646 | 0.406 | 0.157 | 0.430 |
|  | Euclidean global | 0.361 | 0.542 | 0.309 | 0.107 | 0.486 |
|  | Proposed global | 0.247 | 0.378 | 0.116 | 0.115 | 0.380 |
| 95% coverage | Euclidean local | 0.866 | 0.819 | 0.956 | 0.978 | 0.712 |
|  | Euclidean global | 0.935 | 0.893 | 0.957 | 0.980 | 0.911 |
|  | Proposed global | 0.958 | 0.923 | 0.972 | 0.989 | 0.949 |

**Table S3:** Accuracy of Euclidean distance and our proposed distance when using RABC to estimate the four parameters of the global model. For simplicity, we consider a small world of just three countries with different population sizes; here each country’s contribution to the overall accuracy is weighted based on the size of its population.

| Accuracy | Estimation procedure | Average | alpha | beta | delta | gamma |
| --- | --- | --- | --- | --- | --- | --- |
| Absolute bias | Euclidean local | 0.380 | 0.939 | 0.095 | 0.048 | 0.438 |
|  | Euclidean global | 0.194 | 0.338 | 0.064 | 0.025 | 0.351 |
|  | Proposed global | 0.155 | 0.372 | 0.019 | 0.043 | 0.186 |
| Absolute relative bias | Euclidean local | 0.862 | 1.277 | 0.162 | 0.636 | 1.371 |
|  | Euclidean global | 0.512 | 0.459 | 0.101 | 0.304 | 1.185 |
|  | Proposed global | 0.414 | 0.583 | 0.028 | 0.564 | 0.480 |
| IQR range | Euclidean local | 0.147 | 0.228 | 0.156 | 0.048 | 0.154 |
|  | Euclidean global | 0.131 | 0.191 | 0.118 | 0.035 | 0.182 |
|  | Proposed global | 0.073 | 0.111 | 0.033 | 0.030 | 0.120 |
| IQR coverage | Euclidean local | 0.526 | 0.468 | 0.575 | 0.661 | 0.400 |
|  | Euclidean global | 0.610 | 0.548 | 0.641 | 0.701 | 0.550 |
|  | Proposed global | 0.613 | 0.536 | 0.595 | 0.733 | 0.586 |
| 95% range | Euclidean local | 0.360 | 0.571 | 0.361 | 0.128 | 0.380 |
|  | Euclidean global | 0.325 | 0.488 | 0.284 | 0.094 | 0.433 |
|  | Proposed global | 0.201 | 0.310 | 0.094 | 0.090 | 0.309 |
| 95% coverage | Euclidean local | 0.860 | 0.812 | 0.950 | 0.973 | 0.704 |
|  | Euclidean global | 0.928 | 0.887 | 0.954 | 0.976 | 0.898 |
|  | Proposed global | 0.948 | 0.905 | 0.962 | 0.985 | 0.941 |

#### 2.3 Simulation 3: Effectiveness of travel regulation

In this simulation study, we study the effectiveness of different travel regulation policies. We compare the percentages of people allowed to travel under each policy and the pandemic situation in the country adopting the policy. The simulation is set up as follows.

*Step 1. Generating data and parameters*: For *i* ∈ {1, …, *N*} (*N* large), we generate the parameter 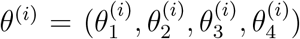, where for each *j* in 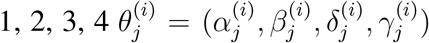, from uniform priors as 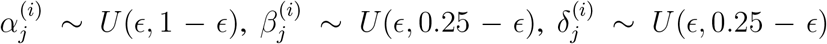, and 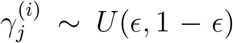. We chose *ϵ* = 0.001 to make sure that the generated parameters do not fall at the boundaries of the priors and cause the generation of atypical data. We also added some constraints to ensure the parameter values are reasonable by only keeping parameters with 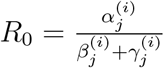 between 0.47 and 6.47 as reported for different regions around the world (*11*). To investigate the effectiveness of travel regulations, we use one more constraint to set the reproduction number *R*_0_ in these 4 countries in 4 different zones, where country 1 has *R*_0_ between 0.47 and 0.9, country 2 has *R*_0_ between 0.9 and 1, country 3 has *R*_0_ between 1 and 1.1, and country 4 has *R*_0_ between 1.1 and 6.47. The initial conditions of each country are generated randomly as 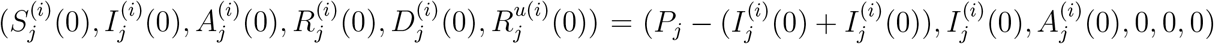, where 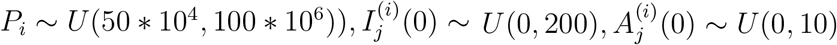. Based on the parameters and the stochastic model, we generate a data set Data^(*i*)^ corresponding to *θ*^(*i*)^. If the generated data set Data^(*i*)^ satisfies the conditions described for Simulation 1 above, we keep *θ*^(*i*)^ and treat it as the underlying true value of the parameter; we also retain the data 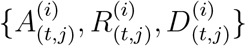 for *j* = 1, 2, 3, 4, which we treat as the observed data collected from each country. We keep generating data till we get 200 underlying true parameters *θ*^(*i*)^ and the corresponding 200 data sets 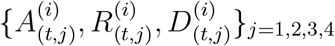. We fix the duration of the simulation to *T* = 42 days for all *i*. Each day, the total number of of outbound travelers from country *j, j* = 1, 2, 3, is drawn from a normal distribution with mean 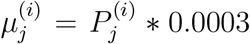 and standard deviation 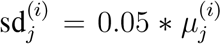, where 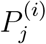 is the size of the population of country *j*. The outbound travelers enter other countries in proportion to sizes of their populations.

*Step 2. Estimation step*: For each iteration *i, i* ∈ {1, …, 200}, based on the sequence of 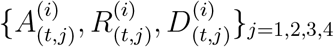, we use the proposed global approach to estimate the underlying true *θ*^(*i*)^. Then 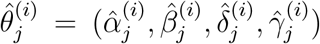 are obtained as the median values of the RABC posterior samples and are used to estimate the underlying true 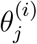, for *j* = 1, 2, 3, 4.

*Step 3. Prediction step*: For each iteration *i, i* ∈ {1, …, 200}, based on the estimated parameter 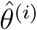, we simulate data for the following two weeks under eight different travel regulation policies. The first two are the most extreme, where all countries are either fully open or fully closed. The third and the fourth are currently used policies, where a 14-days quarantine is required for all arrivals or a 14-day quarantine is required required for arrivals from high-risk countries only. The last 4 policies are our proposed travel regulation policies. We describe each policy in detail below.

P-1 All countries are fully open and allow all airline travel as usual.

P-2 All countries are fully closed and no airline travel is allowed across their borders.

P-3 The country requires a 14-day quarantine for all arrivals. This policy is currently used in many countries such as Korea or India. The other countries are fully open.

P-4 The country requires a 14-day quarantine for travelers from high-risk countries only, i.e., countries with the average number of active confirmed daily cases greater than 20 in 100000 people during the last 2 weeks, and no quarantine for arrivals from other countries. P-4 is a more flexible policy that is currently used by the UK. The other countries are fully open.

P-5 The country adopts a simplified version of the proposed average control policy: we regulate travel such that the average number of daily undetected infected cases is at most 10% higher than the maximum number of daily cases under P-2. The other countries are fully open.

P-6 The country adopts the proposed probability control policy: we regulate travel such that the average number of daily undetected infected cases is at most 10% higher than the maximum number of daily cases under P-2 with probability at least 90%. The other countries are fully open.

P-7 Policy 7 is similar to P-5 but we use the full version of the proposed average control policy as in Example 1 of Section 1.3. The other countries are fully open.

P-8 Policy 8 is similar to P-6 but we use the full version of the proposed probability control policy as in Example 1 of Section 1.3. The other countries are fully open.

Policy effectiveness is evaluated based on two factors: the percentage of people allowed to travel and the pandemic situation in the country once the policy is adopted.

1. The percentage of people allowed to enter the country under each policy is denoted Tc. This number is calculated using the number of people allowed to travel inbound to the country divided by the total number of people willing to enter the country.
2. The percentage of people that will travel due to each policy is denoted Te. This number is an adjusted version Tc. If a 14-day quarantine is applied to a country, we assume that only 5% of the normal number of travelers from this country are willing to travel under this policy. The choice of 5% is based on the data provided by Korea Tourism Organization. (Korea is one of the countries that require a 14-day quarantine for all arrivals.) This adjustment gives us more insights concerning the effect of the 14-day quarantine requirement. After this adjustment, the percentage of expected inbound travelers is obtained by using the number of expected inbound travelers divided by the normal number of inbound travelers.

The effectiveness of policies on the epidemic in the considered country is evaluated based on 7 factors.

1. Percentage of active confirmed imported cases that enter the country due to each policy. This number is calculated using the total number of inbound traveling active confirmed cases that eventually become active confirmed cases, divided by the total number of inbound travelers during the regulation period. We denote this category as IA.
2. Percentage of undetected infected imported cases entering the country due to each policy. This number is obtained using the total number of undetected infected cases traveling inbound divided by the total number of inbound travelers during the regulation period. We denote this category as II.
3. Percentage of undetected infected imported cases when quarantining after entering the country. A policy that does not require quarantine is equivalent to a 0-day quarantine. This number is obtained by taking the total number of undetected infected inbound travelers after quarantine divided by the total number of inbound travelers during the regulation period. We denote this category as IIQ.
4. Relative change in total new cases (detected and undetected), denoted as RU. This number is calculated as the difference in the total number of cases at the end of the regulation period and the beginning of the regulation period, divided by the total number of cases at the beginning of the regulation period.
5. Relative change in total new active confirmed cases, denoted as RA. This number is calculated similarly to RU above but instead of using the number of cases, all counts are based on the number of active confirmed cases.
6. Percent change in total new cases, denoted as PU. This number is calculated as the difference in the total number of cases at the end of the regulation period and the beginning of the regulation period, divided by the population of the country.
7. Percent change in total confirmed cases, denoted as PA. This number is calculated as the difference in the total number of confirmed cases at the end of the regulation period and the beginning of the regulation period, divided by the population of the country.

We generate 1000 stochastic realizations conditional on the estimated parameters and the estimated initial conditions at the beginning of the regulation period. For each realization, we calculate the above metrics, and we report the 0.025 and 0.975 percentile values of each based on the 1000 realizations. To give a fair judgment on the effectiveness of travel regulation on the pandemic, we stratify the metrics by dividing countries to three different groups, where Group 1 corresponds to countries with an effective reproduction number *R*(*t*) lower than 0.9, group 2 corresponds to countries with *R*(*t*) between 0.9 and 1.1, and Group 3 for countries with an *R*_0_ greater than 1.1. Notice that for our model, following Diekmann et al. (2009) (*12*), we can show that the effective reproduction number *R*_*i*_(*t*) of a given country *i* is 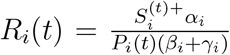. The overall average across these 200 iterations of the above metrics for each group of countries is calculated and used as the final measurement to compare the effectiveness of different policies.

In Table S5, we see that under P-4 the number of expected inbound travelers, Te, is higher than P-5, the simplified version of our proposed average control policy. However, under P-4, the percent of undetected infected after done with quarantine, IIQ enter countries of Group 1 is about (0.03%, 0.03%) and Group 2 is about (0.03%, 0.04%). These values quite high compare to (0.00%, 0.00%) for both groups as in P-5. This is because the average of increased cases each day in the last 14 days was used to decide which countries belong to a green zone or red zone. However, the number of undetected infectious cases may grow very fast in the green zone countries, and in the absence of quarantine, undetected infectious cases from green zone countries may spread the disease fast in the arrival country.

#### 2.4 Simulation 4. Effectiveness of policy coordination

In this simulation study, we study the effectiveness of a global response on the pandemic in terms of the percentage of people allowed to travel and the overall worldwide pandemic situation under a coordinated policy. Simulations are set up similarly to those in Section 2.3 but here we consider a world of 8 countries, where countries 1 and 2 with *R*(0) greater between 1.1 and 6.47, countries 3 and 4 with *R*(0) between 1 and 1.1, countries 5 and 6 with *R*(0) between 0.9 and 1, and countries 7 and 8 with *R*(0) from 0.47 to 0.9.

We consider 8 different policy coordination scenarios:

S-1 All countries are fully open and allow all travel.

S-2 All countries are fully closed and do not allow any airline travel.

S-3 All countries require a 14-day quarantine for all arrivals.

S-4 All countries use the simplified version of the proposed average control policy.

S-5 Countries 1, 3, 5, 7 require a 14-day quarantine for all arrivals, and countries 2, 4, 6, 8 allow no inbound travel.

S-6 Countries 1, 3, 5, 7 use the simplified version of the proposed average control policy, and countries 2, 4, 6, 8 allow no inbound travel.

S-7 Countries 1, 3, 5, 7 require a 14-day quarantine for all arrivals, and countries 2, 4, 6, 8 are fully open.

S-8 Countries 1, 3, 5, 7 use the simplified version of the proposed average control policy, and countries 2, 4, 6, 8 are fully open.

The coordination effectiveness is evaluated based on the overall change in the global pandemic and for each group of countries as in the simulation studies of Section 2.3.

**Table S4:**
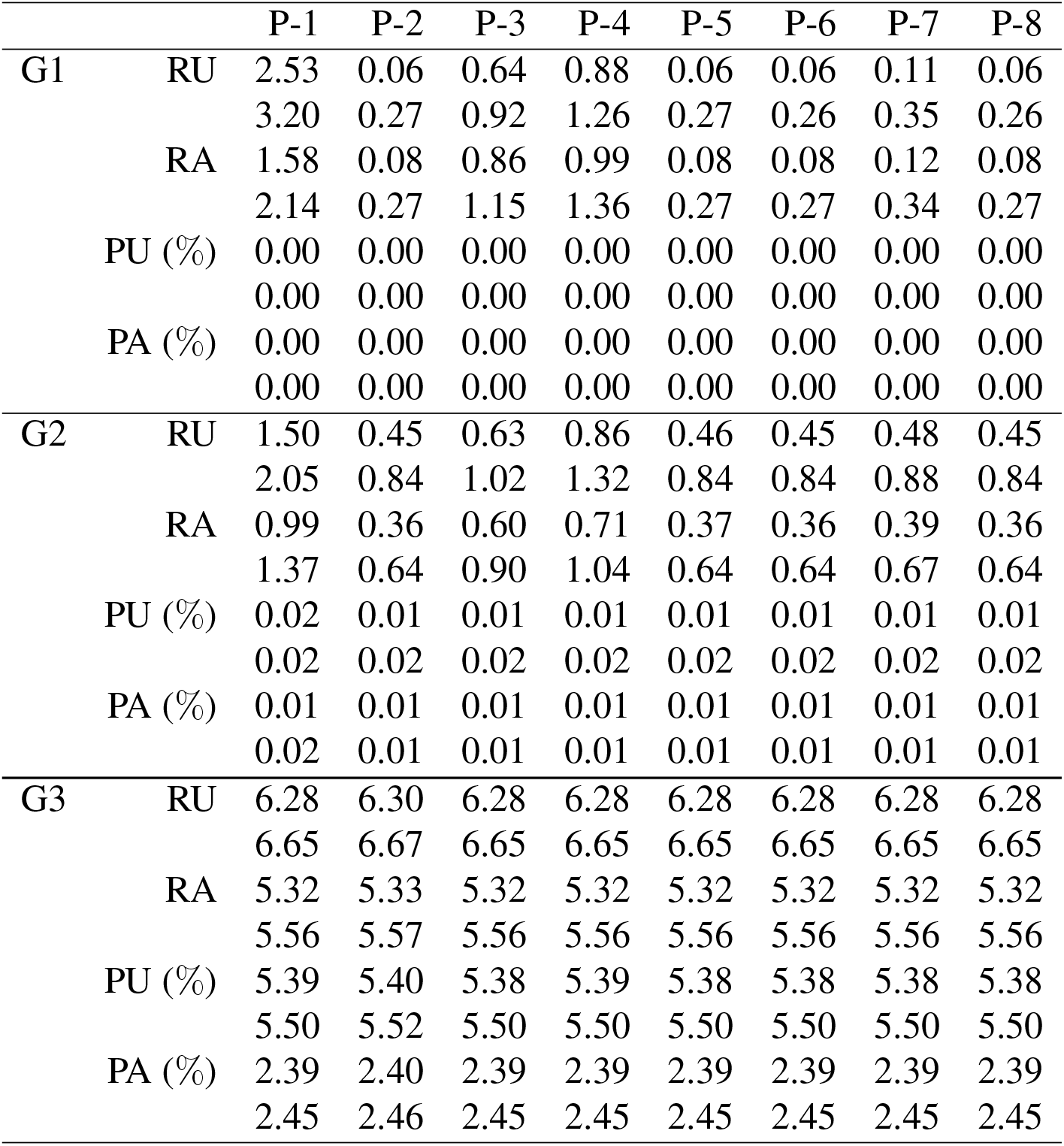
We show (0.025, 0.975) percentiles of pandemic changes for different scenarios. For a given policy, the upper value and lower value of each measurement are the 0.025 percentile value and the 0.975 percentile value, respectively. G1, G2, G3 denotes countries in Group 1, 2, and 3, respectively. RU is the relative change in number of cases (including detected and undetected), RA is the relative change in number of cases that were confirmed, PU is the percent change in total new cases, and PA is the percent change in total new confirmed cases.

**Table S5:** We show (0.025, 0.975) percentiles of travel effects for different policies. For a given policy, the upper value and lower value of each measurement are the 0.025 percentile value and the 0.975 percentile value, respectively. IA is the percentage among the incoming travellers that will eventually become active confirmed after arrival, II is the percentage among the incoming travellers that are undetected infectious, IIQ is the percentage of the incoming travellers who are undetected infectious after the quarantine if the destination country requires a 14-day quarantine, Tc is the percentage of inbound travel capacity, and Te is the percentage of expected of inbound travel.

|  |  | P-1 | P-2 | P-3 | P-4 | P-5 | P-6 | P-7 | P-8 |
| --- | --- | --- | --- | --- | --- | --- | --- | --- | --- |
| G1 | IA (%) | 0.09 | 0.00 | 0.09 | 0.09 | 0.00 | 0.00 | 0.00 | 0.00 |
|  |  | 0.11 | 0.00 | 0.11 | 0.11 | 0.00 | 0.00 | 0.01 | 0.00 |
|  | II (%) | 0.17 | 0.00 | 0.17 | 0.17 | 0.00 | 0.00 | 0.00 | 0.00 |
|  |  | 0.19 | 0.00 | 0.19 | 0.19 | 0.00 | 0.00 | 0.01 | 0.00 |
|  | IIQ (%) | 0.17 | 0.00 | 0.00 | 0.03 | 0.00 | 0.00 | 0.00 | 0.00 |
|  |  | 0.19 | 0.00 | 0.00 | 0.03 | 0.00 | 0.00 | 0.01 | 0.00 |
|  | Tc | 100% | 0% | 100% | 100% | 34% | 0% | 37% | 0% |
|  | Te | 100% | 0% | 5% | 89% | 34% | 0% | 37% | 0% |
| G2 | IA (%) | 0.09 | 0.00 | 0.09 | 0.09 | 0.00 | 0.00 | 0.00 | 0.00 |
|  |  | 0.11 | 0.00 | 0.11 | 0.11 | 0.00 | 0.00 | 0.01 | 0.00 |
|  | II (%) | 0.18 | 0.00 | 0.18 | 0.18 | 0.00 | 0.00 | 0.01 | 0.00 |
|  |  | 0.20 | 0.00 | 0.20 | 0.20 | 0.00 | 0.00 | 0.02 | 0.00 |
|  | IIQ (%) | 0.18 | 0.00 | 0.00 | 0.03 | 0.00 | 0.00 | 0.01 | 0.00 |
|  |  | 0.20 | 0.00 | 0.00 | 0.04 | 0.00 | 0.00 | 0.02 | 0.00 |
|  | Tc | 100% | 0% | 100% | 100% | 60% | 0% | 63% | 0% |
|  | Te | 100% | 0% | 5% | 89% | 60% | 0% | 63% | 0% |
| G3 | IA (%) | 0.00 | 0.00 | 0.00 | 0.00 | 0.00 | 0.00 | 0.00 | 0.00 |
|  |  | 0.00 | 0.00 | 0.00 | 0.00 | 0.00 | 0.00 | 0.00 | 0.00 |
|  | II (%) | 0.00 | 0.00 | 0.00 | 0.00 | 0.00 | 0.00 | 0.00 | 0.00 |
|  |  | 0.00 | 0.00 | 0.00 | 0.00 | 0.00 | 0.00 | 0.00 | 0.00 |
|  | IIQ (%) | 0.00 | 0.00 | 0.00 | 0.00 | 0.00 | 0.00 | 0.00 | 0.00 |
|  |  | 0.00 | 0.00 | 0.00 | 0.00 | 0.00 | 0.00 | 0.00 | 0.00 |
|  | Tc | 100% | 0% | 100% | 100% | 34% | 0% | 34% | 0% |
|  | Te | 100% | 0% | 5% | 100% | 34% | 0% | 34% | 0% |

**Table S6:** We show (0.025, 0.975) percentile of pandemic changes for different scenarios. For a given scenario, the upper value and lower value of each measurement are the 0.025 percentile value and the 0.975 percentile value, respectively. G denotes all countries. See Table S4 caption for more information.

|  |  | S-1 | S-2 | S-3 | S-4 | S-5 | S-6 | S-7 | S-8 |
| --- | --- | --- | --- | --- | --- | --- | --- | --- | --- |
| G | RU | 10.68 | 2.65 | 4.02 | 2.66 | 3.45 | 2.65 | 6.79 | 6.01 |
|  |  | 11.56 | 3.06 | 4.51 | 3.07 | 3.92 | 3.07 | 7.46 | 6.61 |
|  | RA | 8.13 | 2.77 | 4.92 | 2.77 | 4.08 | 2.77 | 6.34 | 5.04 |
|  |  | 8.89 | 3.13 | 5.43 | 3.14 | 4.55 | 3.13 | 6.97 | 5.56 |
|  | PU (%) | 8.35 | 8.31 | 8.28 | 8.31 | 8.29 | 8.31 | 8.31 | 8.33 |
|  |  | 8.39 | 8.35 | 8.32 | 8.35 | 8.33 | 8.35 | 8.35 | 8.37 |
|  | PA (%) | 4.40 | 4.38 | 4.37 | 4.38 | 4.37 | 4.38 | 4.38 | 4.39 |
|  |  | 4.42 | 4.40 | 4.39 | 4.40 | 4.39 | 4.40 | 4.41 | 4.41 |
| G1 | RU | 11.16 | 0.59 | 3.17 | 0.60 | 1.84 | 0.59 | 7.24 | 6.01 |
|  |  | 12.23 | 0.93 | 3.61 | 0.94 | 2.21 | 0.93 | 7.99 | 6.70 |
|  | RA | 9.01 | 0.74 | 4.42 | 0.75 | 2.52 | 0.74 | 6.62 | 4.85 |
|  |  | 9.95 | 1.06 | 4.90 | 1.07 | 2.91 | 1.07 | 7.33 | 5.48 |
|  | PU (%) | 0.05 | 0.01 | 0.02 | 0.01 | 0.01 | 0.01 | 0.03 | 0.03 |
|  |  | 0.05 | 0.01 | 0.02 | 0.01 | 0.01 | 0.01 | 0.04 | 0.03 |
|  | PA (%) | 0.03 | 0.00 | 0.01 | 0.00 | 0.01 | 0.00 | 0.02 | 0.02 |
|  |  | 0.03 | 0.00 | 0.02 | 0.00 | 0.01 | 0.00 | 0.02 | 0.02 |
| G2 | RU | 12.13 | 1.54 | 2.98 | 1.54 | 2.50 | 1.54 | 6.43 | 5.47 |
|  |  | 13.29 | 2.13 | 3.68 | 2.13 | 3.19 | 2.13 | 7.32 | 6.27 |
|  | RA | 8.14 | 1.62 | 4.08 | 1.62 | 3.33 | 1.62 | 5.79 | 4.08 |
|  |  | 9.14 | 2.13 | 4.81 | 2.13 | 4.04 | 2.13 | 6.64 | 4.74 |
|  | PU (%) | 0.11 | 0.04 | 0.05 | 0.04 | 0.05 | 0.04 | 0.08 | 0.08 |
|  |  | 0.13 | 0.05 | 0.06 | 0.05 | 0.06 | 0.05 | 0.10 | 0.09 |
|  | PA (%) | 0.07 | 0.03 | 0.04 | 0.03 | 0.03 | 0.03 | 0.05 | 0.05 |
|  |  | 0.08 | 0.04 | 0.05 | 0.04 | 0.04 | 0.04 | 0.06 | 0.06 |
| G3 | RU | 7.31 | 6.94 | 6.94 | 6.94 | 6.94 | 6.94 | 7.07 | 7.07 |
|  |  | 7.45 | 7.08 | 7.07 | 7.08 | 7.08 | 7.08 | 7.21 | 7.21 |
|  | RA | 7.25 | 7.10 | 7.12 | 7.09 | 7.11 | 7.10 | 7.18 | 7.17 |
|  |  | 7.35 | 7.20 | 7.22 | 7.20 | 7.21 | 7.20 | 7.29 | 7.27 |
|  | PU (%) | 33.11 | 33.14 | 32.99 | 33.14 | 33.06 | 33.14 | 33.05 | 33.12 |
|  |  | 33.24 | 33.27 | 33.12 | 33.27 | 33.19 | 33.27 | 33.18 | 33.25 |
|  | PA (%) | 17.43 | 17.44 | 17.39 | 17.44 | 17.42 | 17.44 | 17.41 | 17.44 |
|  |  | 17.50 | 17.52 | 17.46 | 17.52 | 17.49 | 17.52 | 17.48 | 17.51 |

**Table S7:** We show (0.025, 0.975) percentiles of travel effects for different scenarios. For a given scenario, the upper value and lower value of each measurement are the 0.025 percentile value and the 0.975 percentile value, respectively. See Table S5 caption for more information.

|  |  | S-1 | S-2 | S-3 | S-4 | S-5 | S-6 | S-7 | S-8 |
| --- | --- | --- | --- | --- | --- | --- | --- | --- | --- |
| G | IA (%) | 1.57 | 0.00 | 1.57 | 0.00 | 0.80 | 0.00 | 1.57 | 0.78 |
|  |  | 1.67 | 0.00 | 1.68 | 0.00 | 0.85 | 0.00 | 1.68 | 0.83 |
|  | II (%) | 2.68 | 0.00 | 2.68 | 0.01 | 1.36 | 0.00 | 2.68 | 1.32 |
|  |  | 2.81 | 0.00 | 2.81 | 0.01 | 1.42 | 0.00 | 2.81 | 1.38 |
|  | IIQ (%) | 2.68 | 0.00 | 0.00 | 0.01 | 0.00 | 0.00 | 1.32 | 1.32 |
|  |  | 2.81 | 0.00 | 0.00 | 0.01 | 0.00 | 0.00 | 1.38 | 1.38 |
|  | Tc | 100% | 0% | 100% | 50% | 50% | 25% | 100% | 75% |
|  | Te | 100% | 0% | 5% | 50% | 3% | 25% | 52% | 75% |
| G1 | IA (%) | 1.98 | 0.00 | 1.98 | 0.00 | 0.99 | 0.00 | 1.98 | 0.99 |
|  |  | 2.09 | 0.00 | 2.09 | 0.01 | 1.04 | 0.00 | 2.10 | 1.05 |
|  | II (%) | 3.10 | 0.00 | 3.10 | 0.01 | 1.57 | 0.00 | 3.10 | 1.54 |
|  |  | 3.24 | 0.00 | 3.24 | 0.01 | 1.63 | 0.00 | 3.24 | 1.61 |
|  | IIQ (%) | 3.10 | 0.00 | 0.00 | 0.01 | 0.00 | 0.00 | 1.53 | 1.54 |
|  |  | 3.24 | 0.00 | 0.00 | 0.01 | 0.00 | 0.00 | 1.60 | 1.61 |
|  | Tc | 100% | 0% | 100% | 64% | 50% | 32% | 100% | 82% |
|  | Te | 100% | 0% | 5% | 64% | 3% | 32% | 52% | 82% |
| G2 | IA (%) | 1.77 | 0.00 | 1.77 | 0.00 | 0.89 | 0.00 | 1.77 | 0.89 |
|  |  | 1.89 | 0.00 | 1.89 | 0.01 | 0.95 | 0.00 | 1.90 | 0.95 |
|  | II (%) | 3.02 | 0.00 | 3.02 | 0.01 | 1.52 | 0.00 | 3.02 | 1.51 |
|  |  | 3.17 | 0.00 | 3.17 | 0.01 | 1.59 | 0.00 | 3.17 | 1.59 |
|  | IIQ (%) | 3.02 | 0.00 | 0.00 | 0.01 | 0.00 | 0.00 | 1.51 | 1.51 |
|  |  | 3.17 | 0.00 | 0.00 | 0.01 | 0.00 | 0.00 | 1.58 | 1.59 |
|  | Tc | 100% | 0% | 100% | 64% | 50% | 32% | 100% | 82% |
|  | Te | 100% | 0% | 5% | 64% | 3% | 32% | 52% | 82% |
| G3 | IA (%) | 0.77 | 0.00 | 0.77 | 0.00 | 0.42 | 0.00 | 0.77 | 0.35 |
|  |  | 0.82 | 0.00 | 0.82 | 0.00 | 0.45 | 0.00 | 0.82 | 0.37 |
|  | II (%) | 1.57 | 0.00 | 1.57 | 0.01 | 0.85 | 0.00 | 1.57 | 0.72 |
|  |  | 1.64 | 0.00 | 1.64 | 0.01 | 0.88 | 0.00 | 1.64 | 0.76 |
|  | IIQ (%) | 1.57 | 0.00 | 0.00 | 0.01 | 0.00 | 0.00 | 0.72 | 0.72 |
|  |  | 1.64 | 0.00 | 0.00 | 0.01 | 0.00 | 0.00 | 0.76 | 0.76 |
|  | Tc | 100% | 0% | 100% | 7% | 50% | 3% | 100% | 53% |
|  | Te | 100% | 0% | 5% | 7% | 3% | 3% | 52% | 53% |

## Notes

### Competing Interest Statement

The authors have declared no competing interest.

## References and Notes

1. https://www.worldometers.info/coronavirus/.

2. https://www.worldbank.org/en/news/press-release/2020/06/08/covid-19-to-plunge-global-economy-into-worst-recession-since-world-war-ii.

3. https://www.unwto.org/news/covid-19-international-tourist-numbers-could-fall-60-80-in-2020.

4. https://www.cnbc.com/2020/10/08/over-40-airlines-have-failed-in-2020-so-far-and-more-are-set-to-come.html.

5. https://www.politico.eu/article/coronavirus-travel-economy-193-european-airports-risk-closure-due-to-crisis-industry-lobby/.

6. https://www.nature.com/articles/d41586-020-03605-6.

7. https://news.un.org/en/story/2020/06/1067432.

8. https://www.statista.com/statistics/1104835/coronavirus-travel-tourism-employment-loss/.

9. A. Adekunle, et al., Australian and New Zealand Journal of Public Health 4, 257 (2020).

10. M. Chinazzi, et al., Science 368, 395 (2020).

11. B. Cooper, et al., PLOS Medicine 3, 0845 (2006).

12. K. Linka, et al., Computer Methods in Biomechanics and Biomedical Engineering 23, 710 (2020).

13. T. D. Hollingsworth, et al., Nature Medicine 12, 497 (2006).

14. B. J. Quilty, et al., BMC Medicine (2020).

15. C. R. Wells, et al., PNAS 117, 7504 (2020).

16. N. A. Errett, et al., Journal of Emergency Management 18, 7 (2020).

17. K. A. Grepin, et al., BMJ Global Health (2021).

18. A. L. P. Mateus, et al., Bull World Health Organ 92, 868 (2014).

19. V. Costantino, et al., International Society of Travel Medicine pp. 1–7 (2020).

20. K. Linka, et al., Computational Mechanics (2020).

21. T. Russell, et al., Lancet Public Health 6, 12 (2021).

22. D. J. Warne, et al., BMC Public Health (2020).

23. C. Drovandi, A. N. Pettitt, Biometrics 67, 225 (2011).

24. https://news.sky.com/story/coronavirus-why-countries-are-added-to-uk-quarantine-list-12061651.

25. https://kto.visitkorea.or.kr/eng/tourismstatics/keyfacts/koreamonthlystatistics.kto.

26. https://github.com/cssegisanddata/covid-19.

27. https://opensky-network.org/.

28. E. Lavezzo, et al., Nature 584, 425 (2020).

29. F. Krammer, V. Simon, Science 368, 1060 (2020).

30. R. W. Peeling, et al., Lancet Infectious Disease 20, 245 (2020).

31. H. W. Hethcote, SIAM Review 42, 599 (2000).

32. https://www.nature.com/articles/d41586-020-03370-6.

33. https://www.nature.com/articles/d41586-021-00728-2.

34. J. Lazarus, et al., Nature Medicine 27, 225 (2021).

35. J. Khubchandani, et al., Journal of Community Health 46, 270 (2021).

36. M. Sallam, Vaccine 9 (2021).

37. M. Schwarzinger, et al., Lancet Public Health 6, 210 (2021).

38. M. A. Beaumont, et al. Genetics 162, 2025 (2002).

39. K. Csillery, et al. Trends in Ecology and Evolution 25, 410 (2012).

40. O. Diekmann, et al. J. R. Soc. Interface 7, 873 2010.

41. A. Ebert, https://github.com/AnthonyEbert/protoABC.

42. M. Gatto, et al. PNAS 117, 10484 (2020).

43. M. V. Kiang, et al. Lancet Infectious Disease, 2021.

44. P. Marjoram, et al. PNAS 100, 15324 (2003).

45. D. Prangle, Bayesian Analysis 12, 289 (2017).

46. S. A. Sisson, et al. PNAS 104, 1760 (2007).

47. T. Toni, et al. J. R. Soc. Interface 6, 187 (2009).

## References and Notes

1. D. J. Warne, et al., BMC Public Health (2020).

2. D. T. Gillespie, The Journal of Chemical Physics 115, 1716 (2001).

3. P. Marjoram, et al., PNAS 100, 15324 (2003).

4. S. A. Sisson, et al., PNAS 104, 1760 (2007).

5. T. Toni, et al., J. R. Soc. Interface 6, 187 (2009).

6. C. Drovandi, A. N. Pettitt, Biometrics 67, 225 (2011).

7. https://github.com/anthonyebert/protoabc.

8. M. A. Beaumont, et al., Genetics 162, 2025 (2002).

9. K. Csilléry, et al., Trends in Ecology and Evolution Vol.25 No.7 25, 410 (2012).

10. D. Prangle, Bayesian Analysis 12, 289 (2017).

11. B. Rahman, et al., Wiley (2020).

12. O. Diekmann, et al., J. R. Soc. Interface 7 (2010).

